# Household determinants of animal and human fecal contamination on floors and hands in northwestern coastal Ecuador

**DOI:** 10.64898/2026.08.04.26359736

**Authors:** Viviana Albán, Kelsey J. Jesser, Aldo Lobos, Javier Gallard-Góngora, April M. Ballard, Gwenyth O. Lee, Joseph N.S. Eisenberg, Erica R. Fuhrmeister, Joan A. Casey, Gabriel Trueba, Christine Fagnant-Sperati, Valerie J. Harwood, Karen Levy, ECoMiD Authorship Group

## Abstract

Household environments in low-resource settings can become contaminated with fecal matter from multiple sources, including humans and domestic animals. Identifying the source of fecal contamination is critical for designing targeted interventions to reduce exposure to enteric pathogens, particularly for young children who bear the majority of the enteric disease burden. Using data from 140 households with children enrolled in the ECoMiD cohort study in northwestern coastal Ecuador, we (1) characterized source-specific fecal contamination in samples from household floors and maternal and child hands, and (2) identified animal-related and Water, Sanitation and Hygiene (WASH) conditions associated with the presence and concentrations of these markers. We used five qPCR-based microbial source tracking (MST) markers to detect fecal contamination from avian (GFD), canine (DG37), swine (Pig2Bac), ruminant (Rum2Bac), and human (HF183) sources. Prevalence ratios (PR) and mean differences comparing the presence/absence and concentration, respectively, of MST markers between households with and without each animal-related or WASH condition were estimated using generalized linear models with Poisson and Gaussian distributions. Animal MST markers tracked strongly with several animal-related conditions, whereas associations between the human MST marker and household demographic and WASH conditions were more limited. Animal ownership (PR 1.53; 95% CI: 1.04-2.26) was associated with higher prevalence of animal MST markers on floors. Households reporting animal feces indoors had higher prevalence of animal MST markers on floors (PR 1.84; 95% CI: 1.17-2.89), and higher concentrations of animal MST markers on child hands (mean difference 0.27 gene copies (gc)/m^2^; 95% CI: 0.12-0.41) and maternal hands (mean difference 0.10 gc/m^2^; CI: 0.02-0.18). Animal feces left unremoved outside the home were associated with higher prevalence of animal MST markers on maternal hands (PR 2.31; 95% CI: 1.11-4.81). Mothers reporting direct contact with animals had higher concentrations of animal MST markers on their children’s hands (mean difference 0.12 gc/m^2^; 95% CI: 0.02-0.21). Higher FECEZ scores, an overall metric for animal exposure, were associated with higher prevalence of animal MST markers on maternal hands (PR 2.94; 95% CI: 1.17-7.40). For human fecal contamination, the presence of *E. coli* on child hands was associated with higher prevalence of human MST markers on floors (PR 1.30; 95% CI: 1.00-1.68). Our findings reinforce household floors and maternal and child hands as key reservoirs of fecal contamination and point to future potential targets worth exploring for interventions in similar high-burden settings.

**GRAPHIC ABSTRACT:** 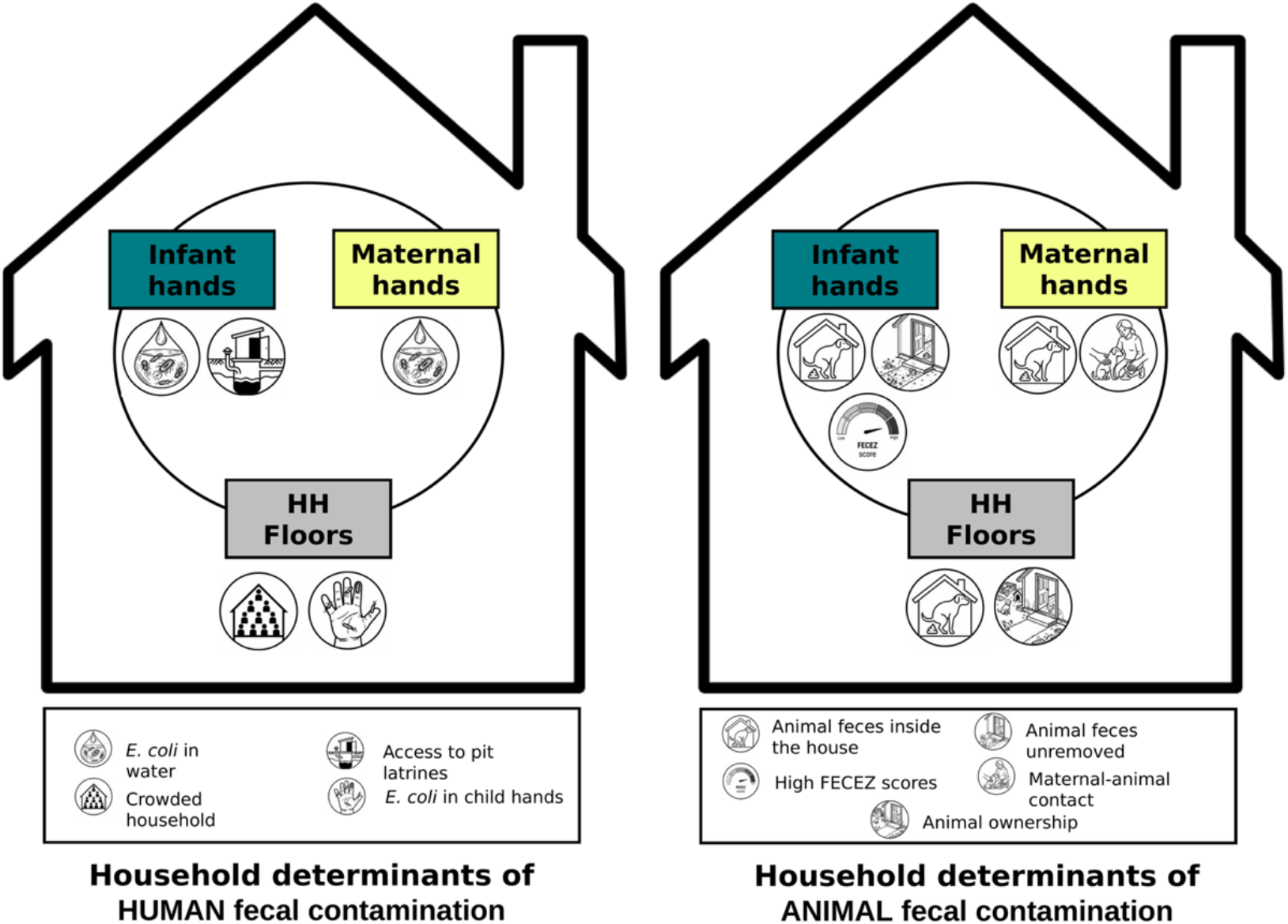

## INTRODUCTION

Household environments in areas with limited access to water, sanitation, and hygiene (WASH) infrastructure, common in many low- and middle-income countries (LMICs), present conditions conducive to transmission of enteric pathogens.[1–3] Fecal material from humans and animals can contaminate diverse household reservoirs, including drinking water sources, food, hands, and fomites, through daily activities and environmental spread, creating multiple opportunities for human exposure to enteric pathogens.[4] Children are especially vulnerable to these exposures, as behaviors like crawling, frequent hand-to-mouth contact, and geophagy[5,6] overlap with immune immaturity, leading to repeated infections and downstream health effects.

Large WASH trials in LMICs have shown mixed effects in reducing child diarrhea and no meaningful improvements on child growth.[7–11] A meta-analysis of several of these WASH trials found only small effects of sanitation interventions on environmental pathogen detection and no effect on the detection of human and animal fecal markers in environmental samples,[12] consistent with the limited observed improvements in child health. These “standard” WASH interventions, which primarily target human excreta, may be insufficient to improve child health because they do not fully address animal feces as a key source of contamination, a factor that may sustain exposure to enteric pathogens. Emerging frameworks such as Transformative WASH[13] and Animal-inclusive Water Sanitation and Hygiene (A-WASH)[14] call for integrating animal husbandry and feces management into child-focused interventions and may be needed to more effectively interrupt transmission in LMIC settings.

Fecal indicator bacteria (FIB), such as *E. coli*, are commonly used to detect fecal contamination but do not distinguish between human and animal sources.[14] Distinguishing animal from human sources of fecal contamination is essential for targeting interventions to reduce environmental transmission of enteric pathogens. Animal fecal contamination may reflect risks tied to husbandry practices and animal feces management in and around the home (e.g., free-roaming animals, accumulation of feces in courtyards or inside homes, disposal practices that spread fecal material onto floors or play areas), while human fecal contamination may point to gaps in sanitation and hygiene (e.g., poorly sealed pits, broken sewer connections, unsafe feces disposal, limited handwashing with soap). Microbial source tracking (MST) offers a useful framework for detecting and quantifying animal versus human sources of fecal contamination, by using quantitative PCR (qPCR) to target gene fragments of enteric bacteria or viruses specific to the feces of a given host species[14,15] from environmental samples.[16]

MST markers have been used in the U.S. primarily to characterize animal and human sources of fecal contamination in recreational waters and to inform water quality management and public health policy.[17,18] In LMICs, MST markers have been applied to differentiate sources of fecal contamination across multiple environmental matrices, including hand rinses, drinking water, soil, and household floors.[19–26] However, important questions remain regarding the relative contribution of animal and human fecal sources to different household reservoirs of microbial contamination, and how source-specific environmental and behavioral factors shape these patterns.

We conducted environmental sampling of floors and child and maternal hands from 140 households enrolled in the ECoMiD birth cohort study in northwestern coastal Ecuador. Five qPCR-based MST markers were used to detect source-specific fecal contamination: avian (GFD), canine (DG37), swine (Pig2Bac), ruminant (Rum2Bac), and human (HF183) markers. These MST markers were selected based on prior validation,[27] and represent the most common animals owned in our study setting.[28,29] Our objectives were to: (i) quantify the prevalence and concentration of human- and animal-associated fecal contamination on household floors and on child and maternal hands using MST; (ii) test if animal exposure was associated with animal-associated MST markers; and (iii) test if household demographics and WASH conditions were associated with human-associated MST markers. This study is unique in concurrently characterizing animal and human fecal sources on household matrices previously identified as key reservoirs of contamination (floors and hands).[27] It also leverages a panel of locally validated MST markers from our study setting[27] and links these source-specific contamination patterns to household-level determinants in a high-burden LMIC setting,[30] providing data useful for the design of potential interventions to reduce the transmission of enteric pathogens.

## METHODS

### Study setting and design

We conducted this study with a subset of households participating in the ECoMiD birth cohort study, (hereafter referred to as the “parent study”), which enrolled 521 pregnant women (at 37 weeks gestation) living in communities along an urban-rural gradient in the province of Esmeraldas, Ecuador. This urban-rural gradient captures a range of environmental exposures. The urban community was represented by the city of Esmeraldas (population ~162,000), the intermediate community by the town of Borbón (population ~5,000) and the rural communities by villages with road access (‘rural-road villages’), including Timbiré, Selva Alegre, Colón Eloy, and Maldonado, and villages accessible only by river (‘rural-river villages’), including Santo Domingo, Zancudo, Colón de Onzole, and San Francisco (rural village populations ~200-1,000 each) (**Figure 1**). The parent study followed children from birth through their first two years of life, collecting time-serial biological samples and surveys at each study visit.[31] In the present study, we measured MST markers in 140 households already enrolled in the parent study. We selected these households based on the child’s age (6, 12, or 18 months at the time of recruitment) and the caregiver’s willingness to participate at the time of recruitment into the MST sub-study. To increase generalizability in our study, we aimed to achieve balance of 06, 12, and 18 months olds across urban, intermediate, and rural communities (either rural-road or rural-river). Due to security concerns in the urban setting during the period of fieldwork, we were unable to reach our intended number of urban households. To compensate and preserve statistical power, we increased the number of participating households from the rural and intermediate communities.

**Figure 1:**
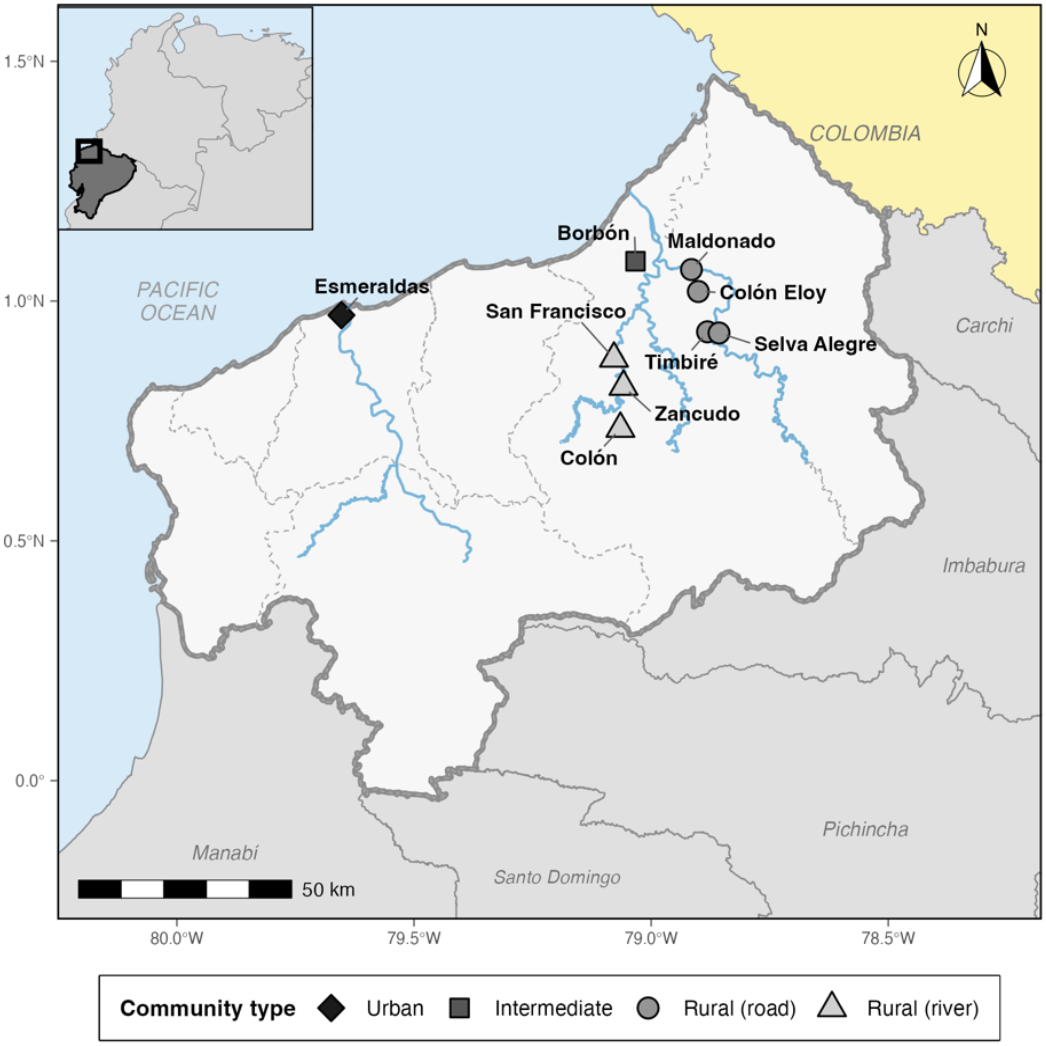
ECoMiD study site map. Map of the nine study communities in Esmeraldas, Province, Ecuador, classified by community type: Urban (n=1), Intermediate (n=1), Rural-road (rural villages with road access) (n=4) and Rural-river (rural villages with river access only) (n=3). Symbols indicate community type. Major rivers are shown in blue. The inset map shows the location of Esmeraldas Province within Ecuador and South America. Scale bar and north arrow are shown in the lower-left and upper-right corners.[32]

### Ethics

All study activities were approved by Institutional Review Boards of the University of Washington (UW; STUDY00014270), Emory University (IRB00101202) and Universidad San Francisco de Quito (IRB2018279 022M and 021-011M), and the Ecuadorian Ministry of Health (MSPCURI000253-4). Participants provided written informed consent prior to survey data and environmental sample collection.

### Data collection

#### Survey data

We used survey data collected by the parent study on household demographics, WASH conditions, animal ownership and animal exposure. Demographic variables were obtained from the baseline survey conducted with pregnant mothers at enrollment. WASH-related variables were drawn from household surveys, and we prioritized data from the survey timepoint that matched the child’s age at the time of recruitment for our sub-study. When data from the same timepoint was not available, we used information from the most recent prior visit to ensure relevance and minimize missing data. A separate survey to gather information on animal-related exposures based on a prior qualitative study[33] was administered to the 140 households in our sub-study concurrently with environmental sampling for MST markers analyses.

#### Environmental sampling and processing

We sampled three matrices - floors, child hands, and maternal hands - guided by prior work in this setting indicating that floors and hands are key reservoirs and exposure pathways for fecal contamination.^35^ All sampling was conducted during the dry season (August–September 2022).

##### Floor samples

We selected floor sampling locations based on maternal reports of where their child spent most of their time. We used a 30 × 30 cm stainless steel surface template, disinfected with 10% bleach followed by 70% ethanol before and after each sampling event. We placed the template on the floor and used two separate sterile cotton swabs for sampling: one pre-moistened with sterile 1X phosphate-buffered saline (PBS), followed by one dry. We passed each swab over the surface within the template area in three directions (vertical, horizontal, and diagonal), to ensure thorough surface coverage, as has been done in previous studies.[22] After sampling, we used sterilized scissors to cut both swabs into a sterile 2 mL microcentrifuge tube containing 1 mL of DNA/RNA Shield buffer (Zymo Research, Irvine, CA) for preservation.

##### Hand rinse samples

We obtained child and maternal hand rinse samples by asking the mothers to insert either their own or their child’s hands into 92 oz sterile Whirl-Pak bags (Filtration Group^®^, Oakbrook Terrace, IL) prefilled with 200 mL of sterile water. While submerged, we gently massaged each hand for 15 seconds and then agitated by shaking the bag for an additional 15 seconds to dislodge any microbial material, as has been done in previous studies.[34] We filtered hand rinse samples by membrane filtration with 47 mm diameter polycarbonate filters (Sigma Millipore) with a pore size of 0.45 μm and flame-sterilized filtration equipment. Following filtration, we stored filtration membranes in sterile 2 mL microcentrifuge tubes containing 1 mL of DNA/RNA Shield buffer to preserve nucleic acid.

##### Field blanks

During each fieldwork day, we collected swab and water field blanks as contamination controls. Sterile swab packaging and Whirl-Pak bags containing sterile water were opened in the household environment for approximately the same amount of time needed for sample collection. Field blank samples were transported alongside unknown samples and processed using the procedures described above.

##### Sample storage and transport

Tubes containing swabs or membrane filters in 1 mL of DNA/RNA Shield buffer were maintained in the field laboratory at ambient temperature for approximately 4 weeks prior to transport to the Universidad San Francisco de Quito (USFQ). Upon arrival, samples were stored at −80°C for approximately 2 weeks until further processing. According to the manufacturer, DNA/RNA Shield buffer preserves nucleic acids at ambient temperature (up to 30°C) for at least 30 days and is stable across a temperature range of −80°C to 70°C, allowing for long-term storage at frozen temperatures.[35]

##### Genomic DNA extraction

At USFQ, we extracted genomic DNA from all samples, including field blanks, using the ZymoBIOMICS DNA Miniprep Kit (Zymo Research) according to the manufacturer’s instructions. Extraction batches consisted of 22 samples, and each batch included an extraction blank with no sample added to monitor potential contamination during processing. DNA was eluted in a total volume of 50 μL and quantified using a Qubit 4 fluorometer (Thermo Fisher Scientific, Waltham, MA). Extracts were stored as two aliquots (20 μL and 30 μL) at −80°C at USFQ. Extracted DNA was later hand-carried to the University of Washington (UW), with cold chain maintained using heavy duty reusable dry ice packs (Techni Ice, Frankston, Australia). DNA was again stored at −80°C upon arrival.

##### MST qPCR assays

Five MST qPCR assays were selected based on their performance in a validation study[27] to detect fecal contamination from specific host sources: human (HF183),[36] avian (GFD),[37] canine (DG37),[38] swine (Pig2Bac),[39] and ruminant (Rum2Bac).[40] Assays for the GFD marker, which uses SYBR-based chemistry, were run in 25 µL reaction volumes with 12.5 µL of Power SYBR Green Master Mix (Thermo Fisher Scientific), 2.5 µL of bovine serum albumin (BSA; Thermo Fisher Scientific) and 2 µL of DNA template per reaction. All other assays rely on TaqMan chemistry and were performed in 25 µL reaction volumes, each containing 12.5 µL of TaqMan™ Environmental Master Mix (Thermo Fisher Scientific) and 2 µL of DNA template. For TaqMan assays, BSA (2.5 µL per reaction) was included in the HF183, DG37, and Pig2Bac assays, but not in the Rum2Bac assay. Primer and probe concentrations for each assay were based on the original protocols described in the corresponding references and are listed in **Table S1**.

We determined MST marker presence and concentration by interpolation against a mean standard curve, calculated from 10 individual standard curves included in each reaction plate. We classified qPCR results as detectable and quantifiable (DQ), detectable but not quantifiable (DNQ), or not detected (ND) based on marker-specific limits of detection (LOD), defined as the lowest amount of template where all triplicate reactions of the standard are detected and limits of quantification (LOQ), defined as the lowest concentration accurately quantified with an acceptable level of uncertainty [CTLOQ = CTLOD – 2(σLOD)]. DQ corresponded to amplification within the standard curve range in both wells, DNQ to amplification below the LOQ but above the LOD, and ND if zero, one, or two wells amplified.[27] Detailed definitions and Ct handling rules are provided in **Table S2**. We used ten-fold serial dilutions of a gBlock gene fragment (Integrated DNA Technologies, Coralville, IA) containing all our MST target sequences to generate six-point standard curves ranging from 50 to 10^6^ gene copies (gc). We ran qPCR reactions for standard curves in triplicate and in duplicate for environmental samples. We evaluated the performance of standard curves following the Minimum Information for Publication of Quantitative Real-Time PCR Experiments (MIQE) guidelines[41] **(Table S3)**. To assess potential qPCR inhibition, we included an inhibition amplification control (IAC) in the HF183 assay according to the recommendations outlined in USEPA Method 1696.[42] No inhibition of qPCR amplification was detected during this study.

### Data analysis

#### Exposure variables

The following exposure variables to characterize the household environment were chosen based on their theoretical relevance to fecal contamination pathways, as informed by the existing literature:[4]

##### Household

We included household crowding (crowded vs. not crowded) and flooring type (improved vs. unimproved). A household was classified as crowded if the ratio of the number of people to the number of rooms exceeded three.[43] Improved flooring was defined as any floor made of cement, ceramic tile, parquet, or polished wood, while unimproved flooring was defined as any floors made of palm/bamboo, earth/sand, or wooden boards. These variables were included as indicators of household socioeconomic status and environmental conditions that may influence exposure to fecal contamination.

##### Urbanicity

Households in the city of Esmeraldas were categorized as “Urban,” households in the town of Borbón as “Intermediate,” and households in Timbiré, Selva Alegre, Colón Eloy, Maldonado, Santo Domingo, Zancudo, Colón de Onzole, and San Francisco as “Rural.” Because the ‘rural-river’ villages included only 12 households, we combined ‘rural-road’ (n=53) and ‘rural-river’ (n=12) villages into a single “Rural” group to ensure as adequate sample size within each urbanicity category.

##### WASH

Water variables included the use of an unimproved drinking water source (unprotected wells, unprotected springs, and surface water from a river or a stream), classified according to the WHO/UNICEF Joint Monitoring Programme (JMP) service ladder for drinking water,[44] and *E. coli* detection in drinking water assessed using the IDEXX method (detected vs. not detected), and drinking water storage practices (yes vs. no). Sanitation was defined based on the household’s toilet discharge system, categorized as flush to sewer (reference group), flush to septic tank, pit latrine, or open defecation/other. Hygiene variables included the detection of *E. coli* on maternal and child hands, assessed using the IDEXX method (presence/absence) and a household hygiene score develop for the parent study. The hygiene score was calculated as the sum of multiple indicators related to kitchen cleanliness, garbage disposal, and handwashing infrastructure, with a possible range of 1–9. Households were classified as less hygienic (scores 1–5) or more hygienic (scores 6–9). Additional details on the construction of this score are described elsewhere.[30]

##### Animal exposures

Animal exposure was the primary exposure of interest and was assessed using the “FECEZ score”, derived from FECEZ Enteropathogens Index, a survey-based metric of fecal-oral transmission of zoonotic enteropathogens developed by our group.[33] We use “FECEZ score” rather than “FECEZ Enteropathogens Index” here because the response format was changed and the principal component analysis used to construct the original index was not repeated. The score comprises 34 items across two domains: child environment and child behavior. Items were adapted from the original instrument by modifying response options from a multi-point Likert scale to binary (yes/no). We calculated a summed score across items and categorized the continuous values into tertiles, expressed as low, medium, or high. Known-groups construct validity of the FECEZ score was assessed following Ballard et al.,[33] recommendations, comparing total and sub-domain scores across community type, child age, and animal ownership; scores were consistently higher in groups with greater expected fecal-oral exposure (rural communities, older children, and households owning animals) (**Figure S1**).

To complement the primary animal exposure variable, we examined additional indicators of animal exposure to capture specific pathways, including ownership, exposures to animal feces, animal feces management, and animal contact. Animal ownership variables included whether the household owned animals (yes vs. no), the number of animals owned (categorized as 0, 1–5, or >5 animals), and whether animals (owned and not owned) were present inside or outside the household (yes vs. no). Variables related to animal feces include the observation of animal feces inside and outside the household (yes vs. no) and animal feces management practices (no visible animal feces present, feces present and removed immediately, feces left sometime before removal or not removed, and other). Animal contact was assessed separately for mothers and children. Contact was defined as engaging in or assisting with any of the following activities: feeding animals, cleaning animal habitats, removing animal feces, or caring for a sick animal.

#### Outcome variables

##### Detection of MST markers

We dichotomized each marker as present (DQ or DNQ) or absent (ND)[41] for each of the three sample types: household floors, child hands, and maternal hands. We also created an “any sample type” variable, defined as the presence of a given marker in at least one of the three sample types to reflect a household-level burden of human fecal contamination. For human fecal contamination, presence/absence was determined using the HF183 marker. For animal fecal contamination, presence/absence was determined using any animal-associated MST marker (GFD, DG37, Rum2Bac, or Pig2Bac). Given the modest sample size and the relatively low prevalence of some individual animal markers (particularly Rum2Bac and Pig2Bac), we *a priori* grouped all animal markers into a combined “Any animal MST” variable to improve statistical power and to capture overall animal fecal contamination from any animal source. Because canine and avian markers were the most prevalent, we also examined them separately in a secondary analysis.

##### Concentration of MST markers

We calculated log-transformed concentrations of MST markers, normalized by sampled surface area, to express all sample types in a common unit area (gc/m^2^). Surface area estimated for hands were based on U.S. EPA-recommended age-specific body part surface areas as outlined in Chapter 7 of the Exposure Factors Handbook (U.S. EPA, 2011): 0.020 m^2^, 0.024, and 0.030 m^2^ for child hands at 6, 12, and 18 months, respectively, and 0.089 m^2^ for maternal hands. Floor sample surface area was standardized to 0.09 m^2^. As with presence/absence, concentration was defined using the HF183 marker for human fecal contamination and as the mean log-transformed concentration across all animal-associated MST markers (GFD, DG37, Rum2Bac and Pig2Bac) for animal fecal contamination, using the same combined “Any animal MST” grouping and for the same reasons described above. We also examined the concentration of canine and avian makers separately as a secondary analysis.

### Statistical analyses

#### Binary analysis (presence/absence of MST markers)

To assess associations between risk factors and the presence of fecal contamination, we used generalized linear models (GLMs) with a Poisson distribution and robust standard errors to estimate prevalence ratios (PRs) and 95% confidence intervals (CIs). Prevalence ratios represent the relative prevalence of MST marker detection in an exposure group compared to a reference group.

#### Continuous analysis (concentration of MST markers)

To assess associations between risk factors and the concentration of fecal contamination, we used GLMs with a Gaussian distribution to estimate mean differences in log-transformed MST marker concentrations and corresponding 95% CIs. Mean differences represent the average change in marker concentration associated with the exposure compared to the reference group.

For both binary and continuous outcomes, we fit separate models for each exposure and outcome pairing to allow for different patterns of fecal contamination, ensure clear interpretation, and reduce concerns about multicollinearity. Multicollinearity was evaluated using variance inflation factors (VIF), with a VIF < 5 considered acceptable. Covariates, encompassing community type (urban, intermediate, rural), socioeconomic status (based on household assets), maternal education, and child’s age were selected *a priori* based on existing literature and a directed acyclic graph (DAG) to identify plausible causal pathways and determine a minimally sufficient adjustment set (**Figure S2**). Community type was included as a covariate in all models, except when community type was the main exposure of interest. All analyses were carried out in R version 4.5.1 using RStudio (v.2026.07.0). GLMs were fitted using the base R glm() function, robust standard errors were calculated using the ‘sandwich’ package, VIFs were assessed using the ‘car’ package, and the directed acyclic graph was constructed using the ‘dagitty’ package.

## RESULTS

### Sample characteristics

The 140 households in our analysis comprised 65 from rural-river and rural-road communities, 55 from the intermediate community, and 20 from the urban community. Most households had access to improved drinking water sources (86.4%), although *E. coli* contamination was detected in 72.1% of drinking water samples. Toilets discharging to septic tanks were the most common sanitation system (42.1%), followed by toilets discharging to sewer system (27.9%), pit latrines (20.0%), and toilets discharging to another place/open defecation (8.6%). On our hygiene scale, 61.4% of households were in the highest hygiene category (scores 6-9), indicating relatively better hygiene practices. Nearly half of the households (48.6%) owned at least one animal, and 13.6% owned more than five animals. Animals were primarily kept outside the household (53.6%), although 20.7% of households reported animals spending time inside the home. Most households observed animal feces in the outdoor household environment (43.6%), and animal feces inside the home was rarely reported (9.3%). We summarize the full distribution of household and WASH-related variables in **Table 1** and animal-related variables in **Table 2**.

**Table 1.** Summary of household and WASH (Water, Sanitation, and Hygiene) variables across communities and all households.

| Household & WASH Variables | Rural* (N=65) | Intermediate (N=55) | Urban (N=20) | All (N=140) |
| --- | --- | --- | --- | --- |
| **Household** |  |  |  |  |
| Improved flooring (%) | 64.6% | 43.6% | 100% | 61.4% |
| Crowded household (%) | 4.6% | 21.8% | 10.0% | 12.1% |
| **Water** |  |  |  |  |
| Improved drinking water source (%) | 76.9% | 92.7% | 100% | 86.4% |
| Stored drinking water (%) | 64.6% | 78.2% | 80.0% | 72.1% |
| *E. coli* detected in drinking water (%)<sup>†</sup> | 96.9% | 69.1% | 50.0% | 79.3% |
| **Hygiene** |  |  |  |  |
| *E. coli* detected on maternal hands (%)<sup>†</sup> | 98.5% | 80.0% | 65.0% | 86.4% |
| *E. coli* detected on child hands (%)<sup>†</sup> | 92.3% | 47.3% | 25.0% | 65.0% |
| Hygiene score (%) |  |  |  |  |
| 1-5 score points | 36.9% | 50.9% | 10.0% | 38.6% |
| 6-9 score points | 63.1% | 49.1% | 90.0% | 61.4% |
| **Sanitation** |  |  |  |  |
| Toilet type (%)<sup>†</sup> |  |  |  |  |
| Flush to sewer | 15.4% | 16.4% | 100% | 27.9% |
| Flush to septic tank | 60.0% | 36.4% | 0% | 42.1% |
| Pit latrine | 2.3% | 36.4% | 0% | 20.0% |
| Flush to another place/Open defecation | 10.8% | 9.1% | 0% | 18.6% |
| Missing | 1.5% | 1.8% | 0% | 1.4% |

\*Rural includes Rural-road and Rural-river communities.
\*Missingness: One household from the urban area had missing data for *E. coli* in drinking water and on maternal and child hands. One rural and one intermediate household were missing data for toilet type

**Table 2.** Summary of animal-related variables across communities and all households.

| **Animal Variables** | **Rural\***<br>**(N=65)** | **Intermediate**<br>**(N=55)** | **Urban**<br>**(N=20)** | **All**<br>**(N=140)** |
| --- | --- | --- | --- | --- |
| **Animal Ownership** |  |  |  |  |
| Animals owned (%) | 58.5% | 49.1% | 15.0% | 48.6% |
| Number of Animals Owned (%) |  |  |  |  |
| 0 animals owned | 41.5% | 50.9% | 85.0% | 51.4% |
| 1-5 animals owned | 40.0% | 40.0% | 5.0% | 35.0% |
| >5 animals owned | 18.5% | 9.1% | 10.0% | 13.6% |
| **Animal Location** |  |  |  |  |
| Animals outside of HH (%) | 63.1% | 58.2% | 10.0% | 53.6% |
| Animals inside of HH (%) | 20.0% | 27.3% | 5.0% | 20.7% |
| **Animal Feces** |  |  |  |  |
| Animal feces outside HH (%) | 44.6% | 52.7% | 15.0% | 43.6% |
| Animal feces inside HH (%) | 9.2% | 9.1% | 10.0% | 9.3% |
| Animal feces (outside) management |  |  |  |  |
| No feces (%) | 55.4% | 47.3% | 85.0% | 56.4% |
| Removed as soon as it was seen (%) | 30.8% | 21.8% | 10.0% | 24.3% |
| Left sometime/ Not removed (%) | 9.2% | 16.4% | 0% | 10.7% |
| Other practice (%) | 4.6% | 14.5% | 5.0% | 8.6% |
| **Animal Contact** |  |  |  |  |
| Child-animal contact (%) | 12.3% | 10.9% | 0% | 10.0% |
| Maternal-animal contact (%) | 27.7% | 45.5% | 0% | 30.7% |
| **FECEZ scores** |  |  |  |  |
| Low scores (%) | 27.7% | 20.0% | 90.0% | 33.6% |
| Medium scores (%) | 40.0% | 34.5% | 10.0% | 33.6% |
| High scores (%) | 32.3% | 45.5% | 0.0% | 32.9% |

\*Rural includes Rural-road and Rural-river communities.

### Household prevalence and concentration of human- and animal-associated MST markers

The human-associated MST marker (HF183) was detected more frequently and at higher concentrations than animal-associated MST markers across all communities and sample types (**Figure 2**).

**Figure 2.**
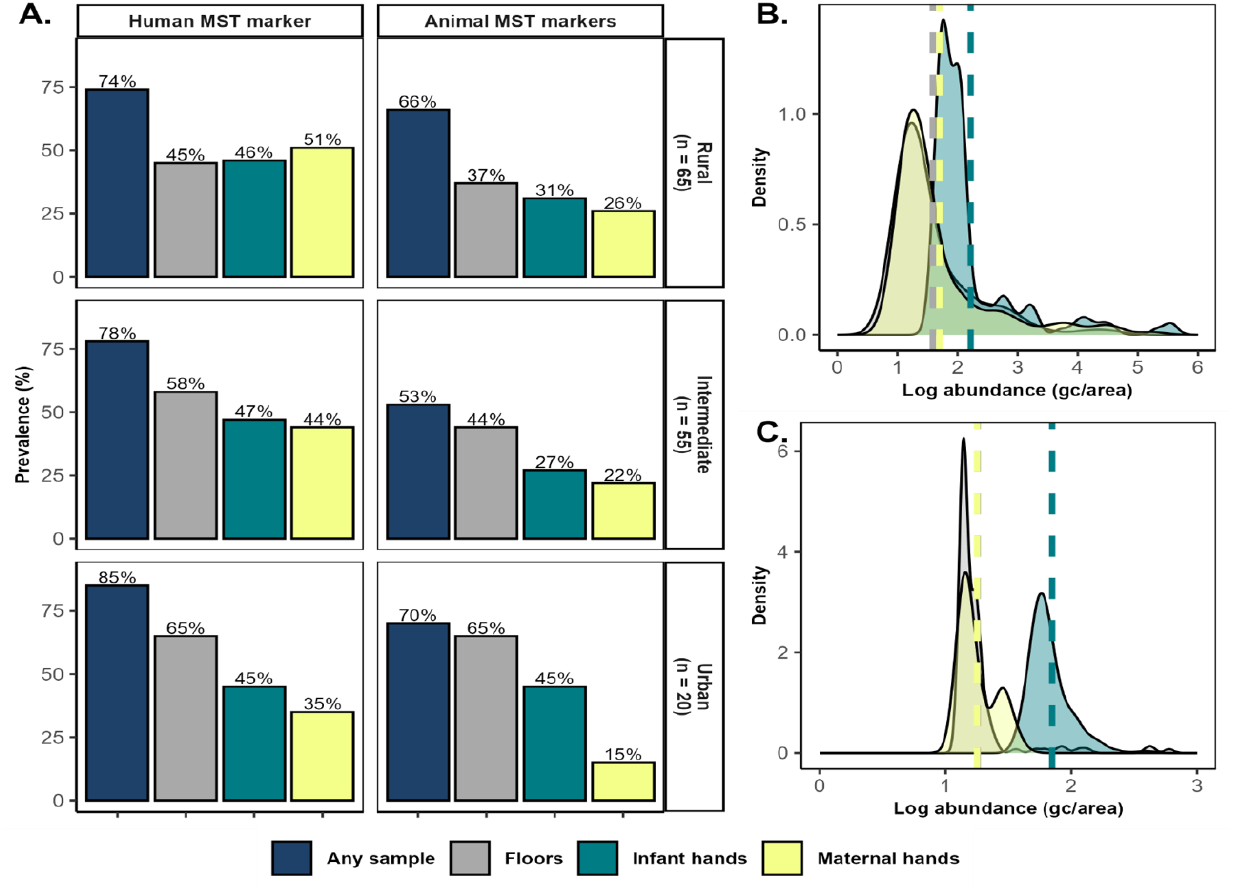
Prevalence and concentration of human and animal Microbial Source Tracking (MST) markers. A. Prevalence of human and animal MST markers by sample type and location. B. Density distribution of the log-transformed abundance (gc/area) of the human-associated MST marker (HF183) across sample types. C. Density distribution of the log-transformed mean abundance (gc/area) of four animal-associated MST markers (GFD, DG37, Pig2Bac and Rum2Bac) across sample types. In density plots (B and C), the y-axis represents the density, where the area under each curve sums to one. Higher peaks indicate a greater concentration of samples around a specific abundance value, while flatter curves indicate more dispersed distributions. Rural includes both Rural-road and Rural-river communities.

At the household level (defined as at least one of the three sample types positive for HF183), 77% of households had evidence of human fecal contamination, while 61% showed evidence of any animal fecal contamination. Floors had higher prevalence of both human (53%) and animal (44%) MST markers, compared to child or maternal hands. However, floors had the lowest mean concentrations of both the human MST marker and animal MST markers. Log-transformed relative concentrations of human MST marker ranged from 1.14 to 5.57 log_10_ gc/m^2^ (**Figure 2B**) and were highest on child hands (2.2 log_10_ gc/m^2^), followed by maternal hands (1.7 log_10_ gc/m^2^) and floors (1.6 log_10_ gc/m^2^). For animal MST markers, log-transformed relative concentrations ranged from 1.14 to 3.67 log_10_ gc/m^2^ (**Figure 2C**), and mean concentrations were also higher on child hands (1.9 log_10_ gc/m^2^), followed by floors (1.3 log_10_ gc/m^2^), and maternal hands (1.3 log_10_ gc/m^2^). The human MST marker was equally prevalent on both child and maternal hands (46% each), whereas animal MST prevalence was higher on child hands (31%) than on maternal hands (23%).

### Household-level determinants of environmental animal and human fecal contamination

We found evidence of associations between several animal-related variables and the prevalence and concentration of animal-associated MST markers. Owning animals was associated with higher animal marker prevalence on floors (PR: 1.53; 95% CI: 1.04, 2.26), with slightly higher prevalence among households owning more than five animals (PR: 1.67; 95% CI: 1.05, 2.65).The presence of animal feces inside the household was associated with animal marker prevalence on floors (PR: 1.84; 95% CI: 1.17, 2.89) and higher concentrations on child hands (Mean diff: 0.27 log_10_gc/m^2^; 95% CI: 0.12, 0.41) and maternal hands (Mean diff: 0.10 log_10_ gc/m^2^; 95% CI: 0.02, 0.18). Animal feces left unremoved outside of the home was also associated with higher prevalence of animal MST markers on maternal hands (PR: 2.31; 95% CI: 1.11–4.81). Maternal contact with animals was associated with higher animal marker concentrations on child hands (Mean diff: 0.12 gc/m^2^; 95% CI: 0.02, 0.21). High FECEZ scores were associated with higher animal marker prevalence on maternal hands (PR: 2.94; 95% CI: 1.17, 7.40) (**Figure 3A-B and Table S4-5)**. In the animal-specific analyses, canine MST marker prevalence on maternal hands was positively associated with animal ownership (particularly 1-5 animals), and with reported presence of animals and their feces inside of the household, while avian MST marker prevalence on floors was positively associated with animal feces outside of the household (**Figure S3**). We did not observe clear associations in canine or avian concentration models (**Figure S4**).

**Figure 3.**
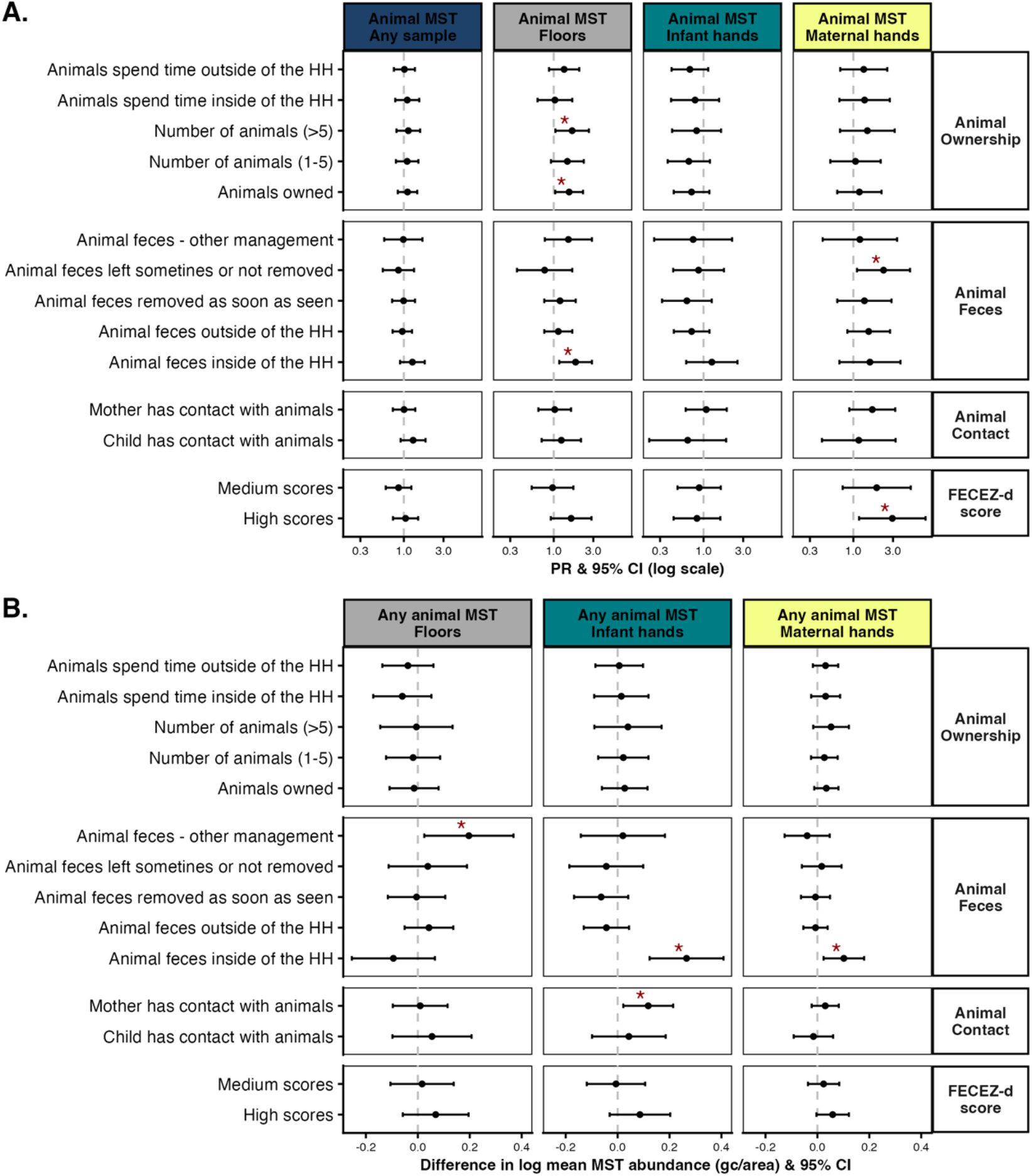
Associations between animal-related factors and environmental animal fecal contamination. **A**. Adjusted prevalence ratios (PRs) and 95% confidence intervals (CIs) for the association between animal ownership, animal feces and animal contacts variables and the presence of any animal-associated microbial source tracking (MST) markers (GFD, DG37, Rum2Bac or Pig2Bac) across different sample types: any sample, floors, child hands, and maternal hands. PRs represent the relative prevalence of MST marker detection in an exposure group compared to a reference group. **B**. Corresponding differences in mean log-transformed of all animal MST present (gc/area) for the same variables and sample types. Statistically significant associations are indicated with a red asterisk (*). Variable domains are grouped by headers on the right. Models were adjusted for community, socio-economic status, maternal education and child’s age.

Evidence for associations between other household-level determinants of environmental human fecal contamination was limited. Detection of *E. coli* in drinking water was associated with higher prevalence of HF183 in any sample (PR: 1.30; 95% CI: 1.00, 1.69). Detection of *E. coli* on child hands was associated with both higher prevalence (PR: 1.53; 95% CI: 1.04, 2.23) and higher concentration (Mean diff: 0.30 log_10_ gc/m^2^; 95% CI: 0.02, 0.57) of HF183 on floors. No meaningful associations were observed for sanitation categories (**Figure 4A-B and Tables S6-7**). In a post hoc analysis prompted by the observation that HF183 prevalence on maternal hands appeared elevated specifically in rural households (**Figure 2A**), we hypothesized that this pattern might be explained by differential WASH access. We tested interaction models between urbanicity and two WASH-related variables, drinking water source and sanitation category for HF183 prevalence on maternal hands. Neither interaction was statistically significant (drinking water source: likelihood ratio test p = 0.39; sanitation category: likelihood ratio test p = 0.79).

**Figure 4.**
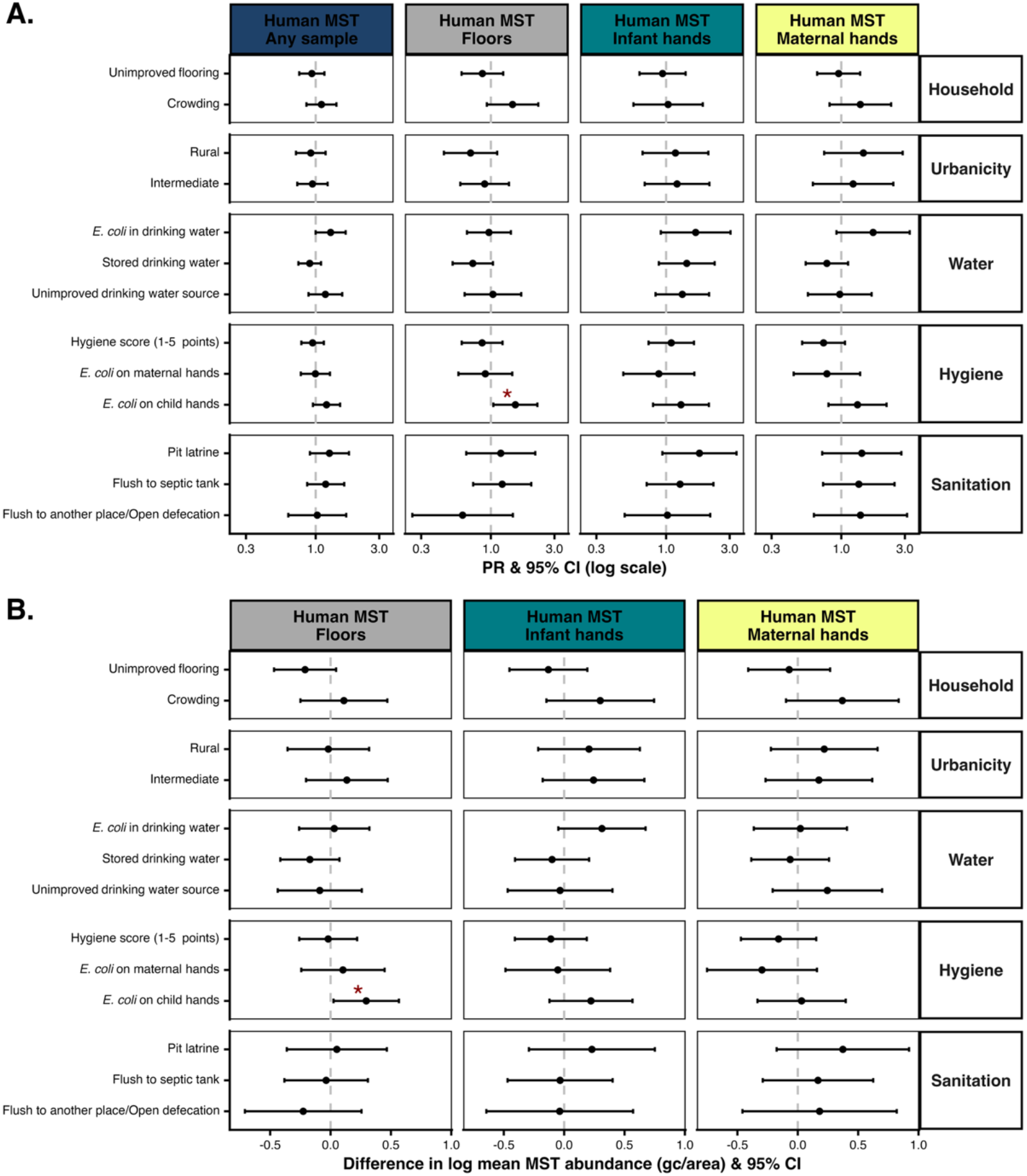
Associations between household and water, sanitation & hygiene (WASH) variables and environmental human fecal contamination. A. Adjusted prevalence ratios (PRs) and 95% confidence intervals (CIs) for the association between household, urbanicity, water, hygiene, and sanitation factors and the presence of the HF183 human-associated microbial source tracking (MST) marker across different sample types: any sample, floors, child hands, and maternal hands. PRs represent the relative prevalence of MST marker detection in an exposure group compared to a reference group. B. Corresponding differences in log-transformed mean HF183 abundance (gc/area) for the same variables and sample types. Associations with statistically significant values are indicated with a red asterisk (*). Variable domains are grouped by headers on the right. Models were adjusted for community, socio-economic status, maternal education and child’s age.

## DISCUSSION

In this study, we show that both human and animal fecal contamination, measured through molecular source-specific MST markers, are ubiquitous in the household environment of children under two years of age in northwestern coastal Ecuador. Animal MST markers tracked strongly with several animal-related conditions, whereas associations between the human MST marker and household demographic and WASH-related conditions were limited. These findings provide evidence on the relative contribution of animal and human fecal sources in the household environment, identify household determinants that may serve as targets for context-specific interventions, particularly for animal fecal contamination in a high-burden LMIC setting, and highlight the utility of animal MST markers to capture animal fecal contamination.

The pattern of low prevalence yet high concentration of animal MST markers, combined with their strong associations with multiple household animal-related conditions, underscores the value of animal MST markers for characterizing animal-sourced fecal contamination and for identifying husbandry practices that generate high-load contamination. The combined “any animal MST” metric captured overall animal fecal contamination, and this aggregated signal tracked strongly with multiple household-level animal conditions. Key determinants of animal fecal contamination in the household environment included the presence of visible animal feces inside the household, ownership of more than five animals, and maternal behaviors related to animal care emerged as key determinants of animal fecal contamination in the household environment.

These findings are consistent with evidence from other LMIC settings, where similar animal-related conditions - animal presence, visible feces, and animal density - have been linked to elevated MST markers, fecal indicator bacteria, and enteric pathogen genes. In rural Bangladesh, the presence of animals and visible animal feces, especially from chickens, sheep/goats, and cows, within the household compound was associated with higher concentrations of indicator *E. coli* on soil floors, and the prevalence of pathogen genes in child stool increased with the number of animal fecal piles in household courtyards.[23,45] Evidence from Peru and Kenya similarly supports the role of animal density and ownership patterns. Households with chickens in Peru had higher odds of detecting avian MST markers (AV4143 and CytB) on floors, and AV4143-positive floors were strongly associated with the presence of *Campylobacter* sp., an important zoonotic enteric pathogen;[21] in urban Kenya, the presence of domestic animals, including chickens, cattle, goats, and sheep, was linked to increased enteric pathogen diversity in public domains, even though concentrations of the indicator enterococci did not differ.[46] Findings from Bangladesh highlight the role of maternal hands in the dynamic transmission of zoonotic pathogens. High concentrations of the animal-associated marker BacCow on maternal hands predicted a greater burden of zoonotic pathogen genes, particularly atypical enteropathogenic *E. coli* (aEPEC), enterohemorrhagic *E. coli* (EHEC), and Shiga toxin-producing *E. coli* (STEC), *and Giardia lamblia*. In addition, aEPEC on mothers’ hands was most strongly associated with cow patties in households.[23] These parallels support the generalizability of our findings and reinforce our interpretation that animal density, feces in and around the home, and maternal animal contact are key animal-related drivers of environmental fecal contamination and child exposure.

Previous qualitative findings from our study area on child play behaviors, animal movement, and feces management practices[29] suggest plausible mechanisms linking animal ownership and visible feces to floor and hand contamination. For example, features of the built environment, such as open doors and lack of fencing, facilitate entry of dogs and free-range chickens into homes where they may defecate. Animal feces observed indoors may reflect not only direct defecation but also tracking of fecal material from outdoor areas on the shoes, feet, or objects carried by household members.[28,29] These mechanisms help explain why floors, more than hands, had the highest prevalence of both human and animal MST marker, despite showing the lowest mean concentration of both markers, consistent with floors functioning as a site of frequent but low-level fecal deposition, rather than a point of concentrated defecation. Together, our MST findings and previous qualitative contextual findings indicate that animal ownership and animal feces in and around the home are consistent markers of conditions that promote the accumulation and spread of animal fecal material on floors and, ultimately, onto maternal and child hands.

Maternal hands emerged as a central interface between animal feces, the broader household environment, and children. The presence of animal feces inside the home, animal feces left unremoved outside of the home, and higher FECEZ scores, were all strongly associated with animal MST markers on maternal hands, and maternal contact with animals was a strong predictor for animal MST markers on child hands. In addition, species-specific secondary analyses supported a particularly important role for maternal hands in canine-associated contamination. These consistent findings highlight the role of mothers and caregivers in mobilizing animal fecal material within the household environment. In our previous study, we found that maternal hands were also a key pathway for human fecal contamination, as detection of the human HF183 marker on maternal hands was associated with its detection on child hands and floors,[27] suggesting that maternal hands may act as a common conduit for both human and animal fecal material. In our study area, mothers commonly care for animals, clean feces, and prepare food and care for children in close succession, providing multiple opportunities for fecal material to be picked up on hands and transferred to floors, utensils, and children.[29] Based on our quantitative associations and the qualitative context, we hypothesize at least two complementary pathways: (1) when animal feces are present inside the home, frequent, close-range contacts lead to high animal marker loads on both maternal and child hands; and (2) when feces remain in peri-domestic spaces, less frequent but repeated encounters during routine movement around the neighborhood result in contamination primarily on maternal hands, which can then be carried indoors. The observed FECEZ score-maternal hand association suggests that as animal exposure accumulates (more animals, more roaming, poorer waste handling), the probability of maternal hand contamination increases, amplifying opportunities to transfer animal fecal material to children during floor play, feeding, and comforting.

The centrality of floors and hands as reservoirs of animal fecal contamination is further supported by and independent study our group conducted in this same area approximately one year earlier (September 2021). Animal MST makers were detected less frequently than human MST markers, but when present, they occurred at high concentrations. This study was designed to validate MST marker performance. It enrolled 58 households selected based on animal ownership rather than drawn from the parent study, and collected more than 10 household environmental samples, including floors and child and maternal hands.[27] In that work, animal MST marker prevalence was 34% on floors, 23% on child hands, and 17% on maternal hands, with concentrations ranging from ~1.5 to 4.0 gc/sample. Despite differences in sampling frame, per-matrix sample size and concentration units that limit precise numerical comparison, the consist pattern of infrequent but high-concentration animal fecal contamination across two independent samples collected roughly a year apart in the same study area supports the robustness of this finding. Results from both studies underscore household floors and hands as key reservoirs and potential transfer points for animal fecal contamination and reinforce the idea that even sporadic animal fecal contamination events may deliver high fecal loads to household reservoirs, on top of the already widespread presence of human fecal contamination.

In contrast to the clear and consistent associations we observed between animal-related variables and animal fecal contamination, determinants of human fecal contamination were much harder to capture. Human fecal contamination was highly prevalent across sample types, with the human HF183 MST marker detected on 53% of floors and 46% of maternal and child hands, often at concentrations similar to or slightly higher than those of animal markers. Despite this widespread contamination, we observed few clear associations between the human MST marker and household demographic and WASH-related indicators. In our post hoc analysis prompted by the elevated prevalence of HF183 on maternal hands in rural households, we found no evidence that differential access to improved water or sanitation explained this pattern, further underscoring their limited explanatory power in this setting. This likely reflects both limitations of standard WASH indicators, which may miss important pathways, such as shared latrines, leaking sewers,[47] unsafe child-feces disposal,[48] and inadequate hand hygiene,[49] and the reality that “improved” infrastructure[50] or reported access does not necessarily translate into effective containment of human excreta.[51] It is also possible that human fecal contamination is so ubiquitous, or so strongly driven by upstream neighborhood-level sanitation and shared infrastructure, that it varies little across households in our study, limiting our ability to identify individual household-level predictors of human fecal contamination. The only meaningful finding was that poor child hand hygiene, measured as presence of indicator *E. coli* on child hands, was associated with the presence and high concentrations of the human MST marker on floors, which is consistent with age-appropriate behaviors such as floor-based play, eating, and drinking;[52–55] however, the cross-sectional nature of our study limits our ability to establish directionality of these relationships.

As a result, our data points to animal-related conditions as potentially more actionable levers for household-level intervention, while also highlighting the need for more sensitive metrics of human sanitation performance. Taken together with prior qualitative work from this setting,[29] which documented multilevel exposure to animal feces, with persistent presence of free-range animals and fecal contamination on child’s play spaces, as well as inadequate practices of animal feces management (e.g., infrequent maternal handwashing after feces removal, inconsistent surface cleaning, and disposal of feces into drains, ditches or surrounding vegetation), our findings reinforce several priorities for further examination. First, limiting animal access indoors to prevent defecation in spaces where children spend time, and promoting safe fecal management practices, for example, using tools to remove feces, cleaning defecation sites with soap and water, and washing hands afterward, are potential household-level strategies. Although our study focused on household determinants, we recognize that exposure to animal feces also occurs outside the home, as described elsewhere.[29,33]

## Strengths and limitations

This study benefits from extensive pre-study validation of both MST assays and field sampling protocols,[27] and from a detailed characterization of animal exposure that moves beyond simple ownership to capture animal husbandry, feces management, and comprehensive exposure assessment through the FECEZ scores.[33] By analyzing both prevalence and concentration of source-specific markers, we provided complementary insights into how often contamination occurs and the magnitude of fecal loads when it does. However, the cross-sectional design limits causal inference and prevents us from establishing directionality of transfer pathways, and our sample size limited our ability to conduct animal species-specific MST models. Future work should use longitudinal, time-aligned designs to resolve directionality and identify high-risk age windows. In addition, some WASH covariates were not measured at the same visit as environmental sampling; this timing mismatch, combined with the inherent measurement noise in WASH indicators, likely introduced nondifferential misclassification, potentially biasing associations toward or away from the null.[56] Our analysis may also be limited by unmeasured confounders, including: parental occupation (e.g., agricultural or livestock work), which could increase household fecal contamination through clothing and footwear; neighborhood-level sanitation conditions that vary beyond the household level; household water chlorination, which may reduce fecal marker detection on hands and could confound associations with WASH-related conditions. These constraints should be considered when interpreting effect sizes and in designing future longitudinal studies.

## Conclusion

Our results are consistent with a growing body of literature linking close contact with domestic animals and their feces to child health risks in LMIC settings, even when conventional WASH infrastructure is present. Studies from rural and peri-urban settings in South Asia and sub-Saharan Africa have reported that free-roaming poultry and livestock, animal corrals adjacent to homes, and animal feces in child play spaces are associated with higher levels of fecal indicator bacteria and enteric pathogens in soil, household surfaces, and children’s stool, as well as with diarrhea and impaired linear growth.[45,52,57,58] Our results reinforce these previous findings that floors and hands, particularly maternal hands, are key reservoirs of animal fecal material. Conditions that increase animal presence and reduce feces management, compound this risk by creating repeated opportunities for children to be exposed to fecal material. By contrast, the ubiquitous nature of human fecal contamination and the limited associations with standard WASH indicators underscore the need for more sensitive metrics (e.g., assessment of shared sanitation infrastructure, monitoring of safe child fecal disposal practices), and to potentially include upstream and community-level drivers of human fecal contamination. These findings, together with the limited impact of large WASH trials that did not explicitly address animal feces, support calls for “transformative WASH”[13] or “Animal-inclusive Water Sanitation and Hygiene (A-WASH)[59] approaches that integrate animal husbandry and feces management into child-focused interventions, rather than treating animals as peripheral to sanitation. Strategies that restrict animal access to child living spaces, promote safe removal and disposal of animal feces, and extend household-level action to shared community environments are worth exploring, to meaningfully reduce child exposure to enteric pathogens in this and similar settings with a high burden of enteric disease.

## Supporting Information

MST qPCR primer and probe sequences and running conditions (Table S1); procedures for determining limits of detection/quantification and marker classification (DQ/DNQ/ND) following MIQE guidelines (Table S2); estimates for associations between animal-related variables and presence/absence and concentration of animal MST markers (Tables S4–S5); estimates for associations between household and WASH-related variables and presence/absence and concentration of the human HF183 MST marker (Tables S6–S7); known-groups construct validity of the FECEZ derived scores (FECEZ scores) (Figure S1); directed acyclic graph of hypothesized relationships among risk factors, covariates, and MST marker outcomes (Figure S2); and canine and avian MST marker associations for presence/absence and concentration (Figures S3–S4) (PDF).

## Data Availability

All data produced in the present study are available upon reasonable request to the authors

## Acknowledgements

This study was funded by the National Institute of Allergy and Infectious Diseases (NIAID) award #R01AI137679 to KL and JNSE, the National Institute of Environmental Health Sciences (NIEHS) awards #T32ES007032-37 and T32ES012870 to KJJ, and the American Association of University Women (AAUW) International Fellowship to VA. The funders had no role in study design, data collection and analysis, decision to publish, or preparation of the manuscript.

