## Supplementary Information for "Household determinants of animal and human fecal contamination on floors and hands in northwestern coastal Ecuador"

### Table of Contents

**Table S1:** MST qPCR reaction primers, probes sequences 3

**Table S2:** Procedures for LOD, LOQ, DQ, DNQ and ND determination following the MIQE guidelines. 4

**Table S4:** Associations between animal-related variables and presence/absences of any animal MST marker 5

**Table S5:** Associations between animal-related variables and mean abundance of all animal MST markers. 6

**Table S6:** Associations between household and WASH-related variables and presence/absences of human HF183 MST marker. 7

**Table S7:** Associations between household and WASH-related variables and abundance of human HF183 MST marker. 8

**Figure S1**: Known groups construct validity of the modified binary FECEZ Enteropathogens 9

**Figure S2.** Directed acyclic graph (DAG). Illustrating the hypothesized relationships between environmental risk factors, covariates, and MST marker presence on floors, child hands, and maternal hands. 10

**Figure S3:** Canine and Avian MST markers (presence/absence) associations 11

**Figure S4:** Canine and Avian MST markers (abundance) associations 12

**Table S1:** MST qPCR reaction primers, probes sequences and running conditions. F: forward; R: reverse; P: probe; IAC: Internal Amplification Control.

| MST marker | Target microorganism | Host | Primer and probe sequences (5'-3') | Running conditions |
|---|---|---|---|---|
| HF183 | (*B. dorei 16S gene*) | Human | F: ATCATGAGTTCACATGTCCG<br>BacR287: CTTCCTCTCAGAACCCCTATCC<br>HF183 P:[FAM]CTAATGGAACGCATCCC[MGB]<br>IAC P: [VIC]AACACGCCGTTGCTACA[MGB] | 10 min at 95°C; 40 cycles of 15 s at 95°C and 1 min at 60°C |
| DG37 | (*Bacteroides*) | Canine | F: TTTTCTCCCACGGTCATCTG<br>R: CTTGGTTATGGGCGACATTG<br>P: [FAM]TGAACGTTTAAAGGAGCAGGTGGCAG[TAMRA] | 15 min at 95°C; 45 cycles of 15 s at 95°C and 1 min at 60°C |
| GFD | (*Helicobacter* spp.) | Avian | F: TCGGCTGAGCACTCTAGGG<br>R: GCGTCTCTTTGTACATCCCA | 10 min at 95°C; 40 cycles of 15 s at 95°C, 30 s at 57°C, and 30 s at 72°C |
| Rum2Bac | (*Bacteroidales*) | Ruminant | F: ACAGCCCGCGATTGATACTGGTAA<br>R: CAATCGGAGTTCTTCGTGAT<br>P: [FAM] ATGAGGTGGATGGAATTCGTGGTGT[BHQ-1] | 10 min at 95°C 40 cycles of 15 s at 95°C and 1 min at 60°C |
| Pig2Bac | (*Bacteroidales*) | Swine | F: GCATGAATTTAGCTTGCTAAATTTGAT<br>R: ACCTCATACGGTATTAATCCGC<br>P: [VIC]TCCACGGGATAGCC [NFQ-MGB] | 10 min at 95°C followed by 40 cycles of 15 s at 95°C and 1 min at 60°C |

**Table S2:** Procedures for LOD, LOQ, DQ, DNQ and ND determination following the MIQE guidelines.

| **qPCR item** | **Description** |
|---|---|
| Limit of Detection (LOD) | The lowest standard on the curve at which at least 2 of 3 replicate reactions amplify. |
| Limit of Quantification (LOQ) | The lowest standard on the curve at which all 3 replicate reactions amplify, with acceptable precision<br>CtLOQ = CtLOD standard – 2σ, where σ is the standard deviation of Ct values across replicate runs of the LOD standard. |
| Detectable Quantifiable (DQ) | Both duplicates amplified, were within 2 Ct values of each other, and fell within the quantifiable range (at or below CtLOQ). The mean of the two duplicate Ct values was assigned. |
| Detectable but not quantifiable (DNQ) | Both duplicates amplified, but above CtLOQ (i.e., between the LOD and LOQ). The Ct value of the LOD standard was assigned. |
| Not detected (ND) | Neither duplicate amplified, or only one of the two amplified. Half the Ct value of the LOD standard was assigned in either case. |

**Table S4:** Associations between animal-related variables and presence/absences of any animal MST marker. Prevalence ratios (PRs), 95% confidence intervals (CIs), and p-values were calculated using a modified Poisson regression model with a log link and robust standard errors. P-values are based on z-statistics computed as the coefficient divided by its robust standard error, assuming a normal distribution.

| Animal-related Variables | Any sample | | | Floors | | | Child hands | | | Maternal hands | | |
|---|---|---|---|---|---|---|---|---|---|---|---|---|
| | PR | [95% CIs] | p-value | PR | [95% CIs] | p-value | PR | [95% CIs] | p-value | PR | [95% CIs] | p-value |
| **Animal Ownership** | | | | | | | | | | | | |
| Animals spend time outside HH<br>***Ref: Animals do not spend time outside*** | 1.02 | [0.76, 1.36] | 0.92 | 1.34 | [0.88, 2.03] | 0.18 | 0.69 | [0.41, 1.14] | 0.14 | 1.33 | [0.70, 2.56] | 0.39 |
| Animals spend time inside HH<br>***Ref: Animals do not spend time outside*** | 1.10 | [0.79, 1.53] | 0.58 | 1.03 | [0.64, 1.67] | 0.89 | 0.79 | [0.41, 1.54] | 0.49 | 1.36 | [0.68, 2.75] | 0.38 |
| Animals owned (>5)<br>***Ref: No animals owned*** | 1.13 | [0.82, 1.57] | 0.46 | 1.67 | [1.05, 2.65] * | 0.03 | 0.83 | [0.42, 1.63] | 0.58 | 1.47 | [0.69, 3.14] | 0.31 |
| Animals owned (1-5)<br>***Ref: No animals owned*** | 1.10 | [0.80, 1.50] | 0.57 | 1.46 | [0.93, 2.30] | 0.10 | 0.67 | [0.37, 1.19] | 0.17 | 1.06 | [0.53, 2.13] | 0.87 |
| Animals owned<br>***Ref: No animals owned*** | 1.11 | [0.85, 1.45] | 0.46 | 1.53 | [1.04, 2.26] * | 0.03 | 0.72 | [0.44, 1.18] | 0.19 | 1.18 | [0.64, 2.18] | 0.59 |
| **Animal Feces** | | | | | | | | | | | | |
| Animal feces-Other management<br>***Ref: No animal feces*** | 0.99 | [0.58, 1.67] | 0.96 | 1.50 | [0.78, 2.89] | 0.22 | 0.75 | [0.25, 2.21] | 0.60 | 1.20 | [0.43, 3.36] | 0.73 |
| Animal feces left sometime/not removed<br>***Ref: No animal feces*** | 0.86 | [0.56, 1.33] | 0.50 | 0.78 | [0.36, 1.67] | 0.52 | 0.87 | [0.43, 1.76] | 0.70 | 2.31 | [1.11, 4.81] * | 0.03 |
| Animal feces removed as soon as seen<br>***Ref: No animal feces*** | 0.99 | [0.72, 1.36] | 0.96 | 1.19 | [0.77, 1.84] | 0.43 | 0.63 | [0.32, 1.26] | 0.19 | 1.36 | [0.64, 2.88] | 0.42 |
| Animal feces outside HH<br>***Ref: No animal feces*** | 0.96 | [0.73, 1.25] | 0.75 | 1.14 | [0.77, 1.67] | 0.52 | 0.72 | [0.44, 1.18] | 0.19 | 1.53 | [0.85, 2.76] | 0.16 |
| Animal feces inside HH<br>***Ref: No animal feces*** | 1.27 | [0.90, 1.78] | 0.17 | 1.84 | [1.17, 2.89] * | 0.01 | 1.26 | [0.62, 2.56] | 0.52 | 1.58 | [0.68, 3.69] | 0.29 |
| **Animal Contact** | | | | | | | | | | | | |
| Maternal-animal contact<br>***Ref: No contact*** | 1.00 | [0.74, 1.37] | 0.98 | 1.03 | [0.65, 1.62] | 0.11 | 1.08 | [0.61, 1.90] | 0.79 | 1.69 | [0.89, 3.19] | 0.90 |
| Child-animal contact<br>***Ref: No contact*** | 1.29 | [0.91, 1.82] | 0.15 | 1.24 | [0.72, 2.13] | 0.45 | 0.65 | [0.22, 1.87] | 0.42 | 1.16 | [0.42, 3.22] | 0.77 |
| **FECEZ-d scores** | | | | | | | | | | | | |
| Medium scores<br>***Ref: Low score*** | 0.86 | [0.61, 1.23] | 0.42 | 0.97 | [0.54, 1.73] | 0.91 | 0.89 | [0.49, 1.61] | 0.69 | 1.91 | [0.74, 4.91] | 0.18 |
| High score<br>***Ref: Low score*** | 1.05 | [0.74, 1.49] | 0.79 | 1.62 | [0.92, 2.85] | 0.09 | 0.83 | [0.44, 1.59] | 0.58 | 2.94 | [1.17, 7.40] * | 0.02 |

**Table S5:** Associations between animal-related variables and mean abundance of all animal MST markers. Mean differences (PRs), 95% confidence intervals (CIs), and p-values were calculated using a generalized linear (Gaussian) regression model. P-values are based on z-statistics computed as the coefficient divided by its robust standard error, assuming a normal distribution.

| Animal-related Variables | Floors | | | Child hands | | | Maternal hands | | |
|---|---|---|---|---|---|---|---|---|---|
| | Mean diff. | [95% CIs] | p-value | Mean diff. | [95% CIs] | p-value | Mean diff. | [95% CIs] | p-value |
| **Animal Ownership** | | | | | | | | | |
| Animals spend time outside HH<br>***Ref: Animals do not spend time outside*** | -0.04 | [-0.14, 0.06] | 0.45 | 0.01 | [-0.09, 0.10] | 0.89 | 0.03 | [-0.02, 0.08] | 0.21 |
| Animals spend time inside HH<br>***Ref: Animals do not spend time outside*** | -0.06 | [-0.17, 0.05] | 0.30 | 0.01 | [-0.09, 0.12] | 0.79 | 0.03 | [-0.02, 0.09] | 0.27 |
| Animals owned (>5)<br>***Ref: No animals owned*** | -0.01 | [-0.14, 0.13] | 0.94 | 0.04 | [-0.09, 0.17] | 0.55 | 0.05 | [-0.02, 0.12] | 0.14 |
| Animals owned (1-5)<br>***Ref: No animals owned*** | -0.02 | [-0.12, 0.09] | 0.73 | 0.02 | [-0.07, 0.12] | 0.66 | 0.03 | [-0.02, 0.08] | 0.31 |
| Animals owned<br>***Ref: No animals owned*** | -0.01 | [-0.11, 0.08] | 0.76 | 0.03 | [-0.06, 0.12] | 0.54 | 0.03 | [-0.01, 0.08] | 0.15 |
| **Animal Feces** | | | | | | | | | |
| Animal feces-Other management<br>***Ref: No animal feces*** | 0.20 | [0.02, 0.37] * | 0.03 | 0.02 | [-0.14, 0.18] | 0.81 | -0.04 | [-0.13, 0.05] | 0.38 |
| Animal feces left sometime/not removed<br>***Ref: No animal feces*** | 0.04 | [-0.11, 0.19] | 0.62 | -0.04 | [-0.19, 0.10] | 0.55 | 0.02 | [-0.06, 0.09] | 0.67 |
| Animal feces removed as soon as seen<br>***Ref: No animal feces*** | 0.00 | [-0.12, 0.11] | 0.93 | -0.06 | [-0.17, 0.04] | 0.24 | -0.01 | [-0.06, 0.05] | 0.80 |
| Animal feces outside HH<br>***Ref: No animal feces*** | 0.04 | [-0.05, 0.14] | 0.37 | -0.04 | [-0.13, 0.04] | 0.33 | -0.01 | [-0.05, 0.04] | 0.76 |
| Animal feces inside HH<br>***Ref: No animal feces*** | -0.09 | [-0.25, 0.07] | 0.25 | 0.27 | [0.12, 0.41] * | 0.00 | 0.10 | [0.02, 0.18] * | 0.01 |
| **Animal Contact** | | | | | | | | | |
| Maternal-animal contact<br>***Ref: No contact*** | 0.01 | [-0.10, 0.11] | 0.87 | 0.12 | [0.02, 0.21] * | 0.02 | 0.03 | [-0.02, 0.08] | 0.26 |
| Child-animal contact<br>***Ref: No contact*** | 0.05 | [-0.10, 0.21] | 0.48 | 0.04 | [-0.10, 0.19] | 0.55 | -0.02 | [-0.09, 0.06] | 0.69 |
| **FECEZ-d scores** | | | | | | | | | |
| Medium scores<br>***Ref: Low score*** | 0.02 | [-0.11, 0.14] | 0.79 | -0.01 | [-0.12, 0.11] | 0.91 | 0.02 | [-0.04, 0.08] | 0.44 |
| High score<br>***Ref: Low score*** | 0.07 | [-0.06, 0.20] | 0.29 | 0.09 | [-0.03, 0.20] | 0.15 | 0.06 | [0.00, 0.12] | 0.07 |

**Table S6:** Associations between household and WASH-related variables and presence/absences of human HF183 MST marker. Prevalence ratios (PRs), 95% confidence intervals (CIs), and p-values were calculated using a modified Poisson regression model with a log link and robust standard errors. P-values are based on z-statistics computed as the coefficient divided by its robust standard error, assuming a normal distribution.

| Household & WASH-related Variables | Any sample | | | Floors | | | Child hands | | | Maternal Hands | | |
|---|---|---|---|---|---|---|---|---|---|---|---|---|
| | PR | [95% CIs] | p-value | PR | [95% CIs] | p-value | PR | [95% Cis] | p-value | PR | [95% Cis] | p-value |
| **Household factors** | | | | | | | | | | | | |
| Unimproved floor<br>***Ref:*** *Improved floor* | 0.94 | [0.75, 1.16] | 0.55 | 0.86 | [0.60, 1.24] | 0.42 | 0.94 | [0.63, 1.40] | 0.76 | 0.96 | [0.66, 1.38] | 0.81 |
| Crowded<br>***Ref:*** *Uncrowded* | 1.10 | [0.85, 1.43] | 0.45 | 1.45 | [0.93, 2.27] | 0.10 | 1.04 | [0.57, 1.89] | 0.91 | 1.39 | [0.82, 2.37] | 0.23 |
| **Urbanicity** | | | | | | | | | | | | |
| Rural<br>***Ref:*** *Urban* | 0.92 | [0.71, 1.19] | 0.52 | 0.70 | [0.44, 1.11] | 0.13 | 1.18 | [0.67, 2.08] | 0.58 | 1.46 | [0.74, 2.89] | 0.27 |
| Intermediate<br>***Ref:*** *Urban* | 0.95 | [0.73, 1.23] | 0.68 | 0.90 | [0.59, 1.37] | 0.62 | 1.21 | [0.69, 2.11] | 0.51 | 1.23 | [0.61, 2.46] | 0.56 |
| **Water** | | | | | | | | | | | | |
| *E. coli* in drinking water<br>***Ref:*** *No E. coli in drinking water* | 1.30 | [1.00, 1.68] * | 0.05 | 0.97 | [0.66, 1.41] | 0.86 | 1.67 | [0.91, 3.04] | 0.10 | 1.73 | [0.92, 3.26] | 0.09 |
| Stored drinking water<br>***Ref:*** *Non stored water* | 0.90 | [0.74, 1.10] | 0.30 | 0.73 | [0.52, 1.04] | 0.08 | 1.43 | [0.88, 2.32] | 0.15 | 0.78 | [0.54, 1.12] | 0.18 |
| Unimproved drinking water source<br>***Ref:*** *Improved water source* | 1.18 | [0.88, 1.58] | 0.26 | 1.04 | [0.64, 1.69] | 0.89 | 1.32 | [0.83, 2.10] | 0.24 | 0.97 | [0.56, 1.69] | 0.93 |
| **Hygiene** | | | | | | | | | | | | |
| Hygiene score (1-5 points)<br>***Ref:*** *Hygiene score (6-9 points)* | 0.95 | [0.78, 1.15] | 0.59 | 0.86 | [0.60, 1.22] | 0.40 | 1.09 | [0.74, 1.62] | 0.65 | 0.74 | [0.51, 1.07] | 0.10 |
| *E. coli* on maternal hands<br>***Ref:*** *Not E. coli in maternal hands* | 1.00 | [0.77, 1.28] | 0.97 | 0.91 | [0.57, 1.45] | 0.69 | 0.88 | [0.48, 1.63] | 0.69 | 0.78 | [0.44, 1.39] | 0.40 |
| *E. coli* on child hands<br>***Ref:*** *Nor E. coli in child hands* | 1.21 | [0.95, 1.53] | 0.12 | 1.53 | [1.04, 2.23] * | 0.03 | 1.29 | [0.80, 2.10] | 0.30 | 1.32 | [0.80, 2.19] | 0.27 |
| **Sanitation** | | | | | | | | | | | | |
| Pit latrine<br>***Ref:*** *Flush to sewer* | 1.27 | [0.91, 1.78] | 0.17 | 1.19 | [0.65, 2.15] | 0.57 | 1.78 | [0.94, 3.38] | 0.08 | 1.42 | [0.72, 2.82] | 0.31 |
| Flush to septic tank<br>***Ref:*** *Flush to sewer* | 1.19 | [0.86, 1.64] | 0.29 | 1.22 | [0.74, 2.01] | 0.44 | 1.27 | [0.72, 2.27] | 0.41 | 1.35 | [0.73, 2.51] | 0.34 |
| Flush to another place/Open defecation<br>***Ref:*** *Flush to sewer* | 1.03 | [0.62, 1.69] | 0.91 | 0.61 | [0.26, 1.46] | 0.27 | 1.02 | [0.49, 2.14] | 0.95 | 1.39 | [0.62, 3.12] | 0.42 |

**Table S7:** Associations between household and WASH-related variables and abundance of human HF183 MST marker. Mean differences (PRs), 95% confidence intervals (CIs), and p-values were calculated using a generalized linear (Gaussian) regression model. P-values are based on z-statistics computed as the coefficient divided by its robust standard error, assuming a normal distribution.

| **Household & WASH-related Variables** | **Floors** | | | **Child hands** | | | **Maternal hands** | | |
|---|---|---|---|---|---|---|---|---|---|
| | **Mean diff.** | **[95% CIs]** | **p-value** | **Mean diff.** | **[95% CIs]** | **p-value** | **Mean diff.** | **[95% CIs]** | **p-value** |
| **Household factors** | | | | | | | | | |
| Unimproved floor<br>***Ref:*** *Improved floor* | -0.21 | [-0.47, 0.05] | 0.11 | -0.13 | [-0.45, 0.19] | 0.43 | -0.07 | [-0.41, 0.27] | 0.68 |
| Crowded<br>***Ref:*** *Uncrowded* | 0.11 | [-0.25, 0.47] | 0.55 | 0.30 | [-0.15, 0.75] | 0.19 | 0.37 | [-0.10, 0.84] | 0.12 |
| **Urbanicity** | | | | | | | | | |
| Rural<br>***Ref:*** *Urban* | -0.02 | [-0.36, 0.32] | 0.92 | 0.21 | [-0.22, 0.63] | 0.34 | 0.22 | [-0.22, 0.66] | 0.33 |
| Intermediate<br>***Ref:*** *Urban* | 0.14 | [-0.20, 0.47] | 0.43 | 0.24 | [-0.18, 0.66] | 0.26 | 0.17 | [-0.27, 0.62] | 0.44 |
| **Water** | | | | | | | | | |
| *E. coli* in drinking water<br>***Ref:*** *No E. coli in drinking water* | 0.03 | [-0.26, 0.32] | 0.84 | 0.31 | [-0.05, 0.67] | 0.09 | 0.02 | [-0.36, 0.41] | 0.91 |
| Stored drinking water<br>***Ref:*** *Non stored water* | -0.17 | [-0.42, 0.07] | 0.17 | -0.10 | [-0.41, 0.21] | 0.52 | -0.06 | [-0.39, 0.26] | 0.70 |
| Unimproved drinking water source<br>***Ref:*** *Improved water source* | -0.09 | [-0.44, 0.26] | 0.62 | -0.03 | [-0.47, 0.40] | 0.88 | 0.24 | [-0.21, 0.70] | 0.29 |
| **Hygiene** | | | | | | | | | |
| Hygiene score (1-5 points)<br>***Ref:*** *Hygiene score (6-9 points)* | -0.02 | [-0.26, 0.22] | 0.87 | -0.11 | [-0.41, 0.19] | 0.47 | -0.16 | [-0.47, 0.15] | 0.32 |
| *E. coli* on maternal hands<br>***Ref:*** *Not E. coli in maternal hands* | 0.10 | [-0.24, 0.45] | 0.56 | -0.05 | [-0.49, 0.38] | 0.81 | -0.30 | [-0.75, 0.16] | 0.20 |
| *E. coli* on child hands<br>***Ref:*** *Nor E. coli in child hands* | 0.30 | [0.02, 0.57]* | 0.03 | 0.22 | [-0.12, 0.57] | 0.21 | 0.03 | [-0.33, 0.40] | 0.87 |
| **Sanitation** | | | | | | | | | |
| Pit latrine<br>***Ref:*** *Flush to sewer* | 0.05 | [-0.36, 0.47] | 0.81 | 0.23 | [-0.29, 0.75] | 0.39 | 0.37 | [-0.18, 0.92] | 0.18 |
| Flush to septic tank<br>***Ref:*** *Flush to sewer* | -0.04 | [-0.38, 0.31] | 0.84 | -0.03 | [-0.47, 0.40] | 0.88 | 0.17 | [-0.29, 0.62] | 0.47 |
| Flush to another place/Open defecation<br>***Ref:*** *Flush to sewer* | -0.23 | [-0.71, 0.26] | 0.36 | -0.04 | [-0.64, 0.57] | 0.91 | 0.18 | [-0.46, 0.82] | 0.58 |

**Figure S1**: Known groups construct validity of the FECEZ-derived scores (FECEZ-d scores). Boxplots show total score (top row), environment sub-domain (middle row), and behavior sub-domain (bottom row) across community type, child age, and household animal ownership. Pairwise comparisons for community type and child age are Tukey-adjusted; animal ownership was compared using the Wilcoxon rank-sum test. ****p<0.0001; *p<0.05.

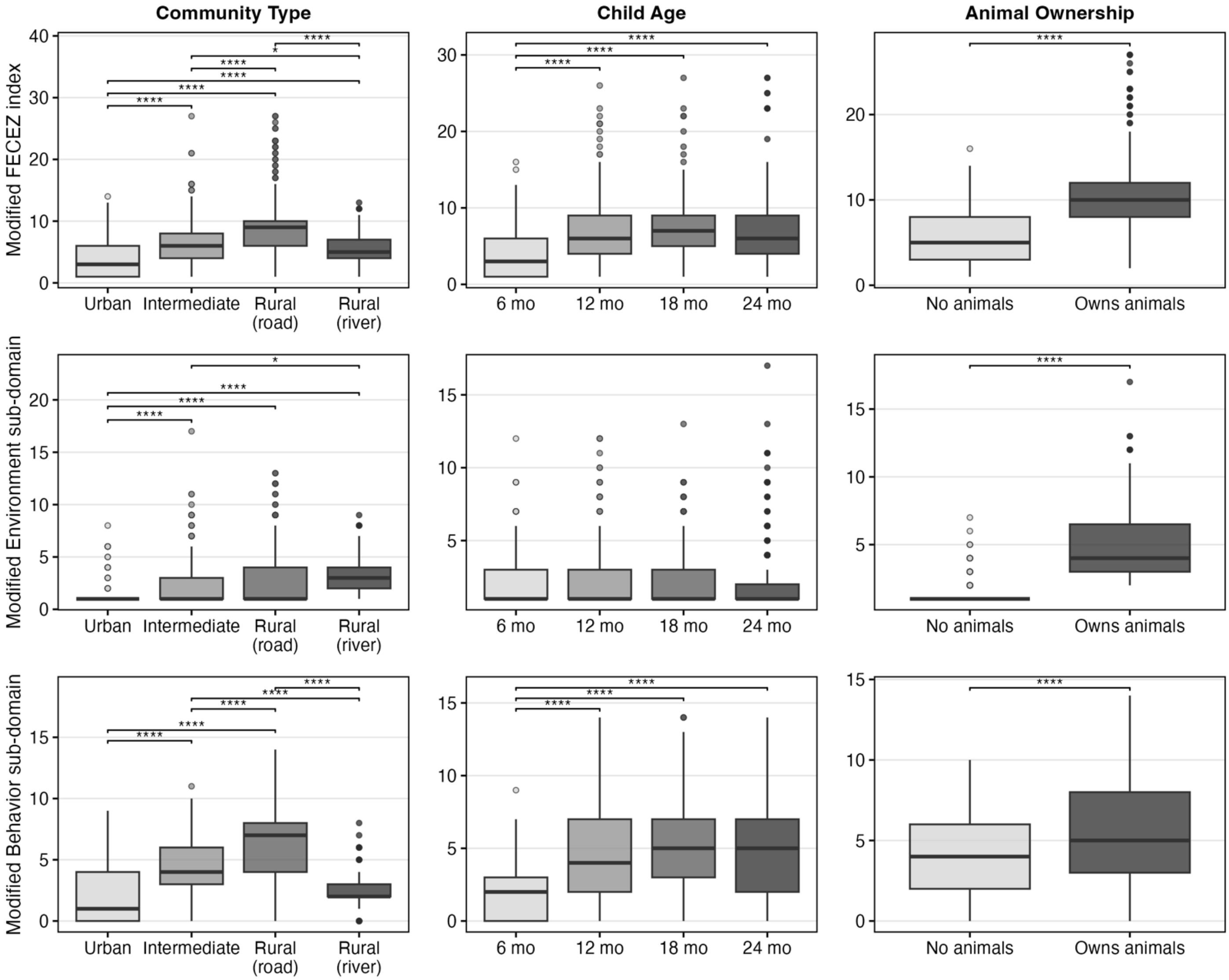

**Figure S2.** Directed acyclic graph (DAG). Illustrating the hypothesized relationships between environmental risk factors, covariates, and MST marker presence on floors, child hands, and maternal hands. The red-highlighted variables represent the minimal sufficient adjustment set required to block all backdoor pathways between exposure and outcome. SES: Socio-economic status.

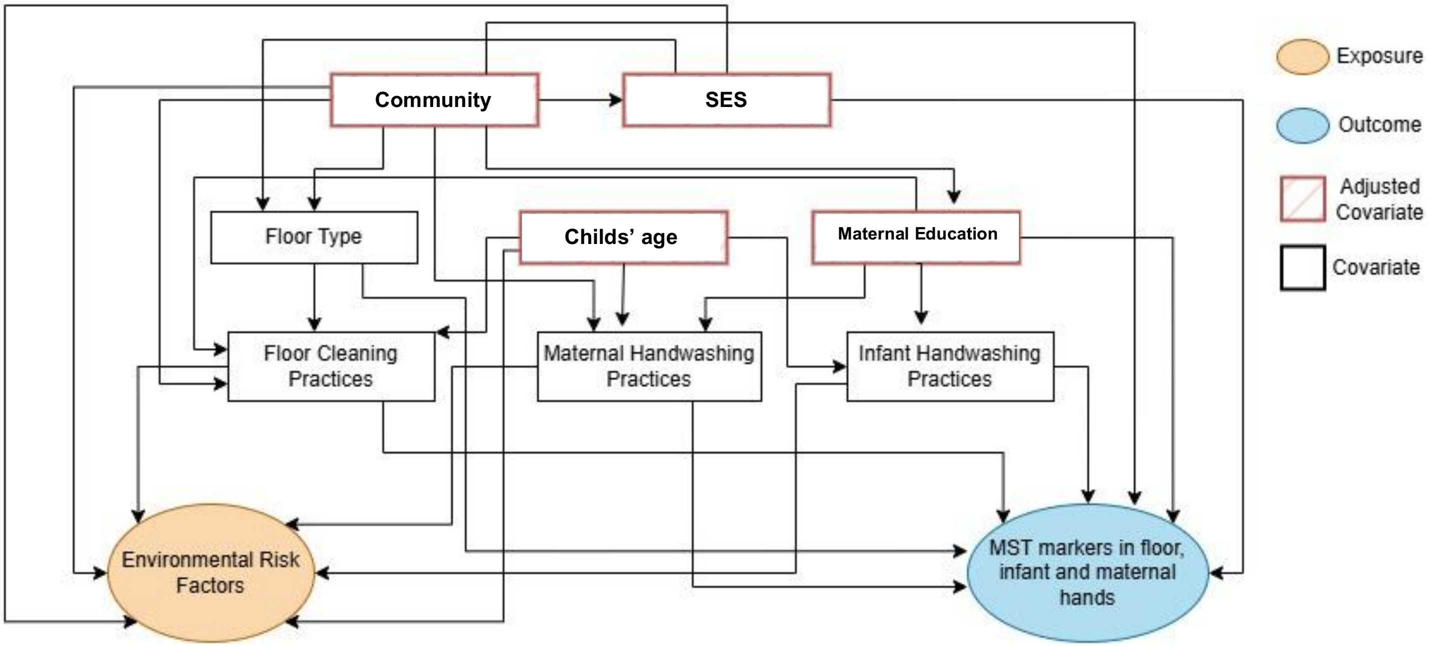

**Figure S3:** Canine and Avian MST markers (presence/absence) associations

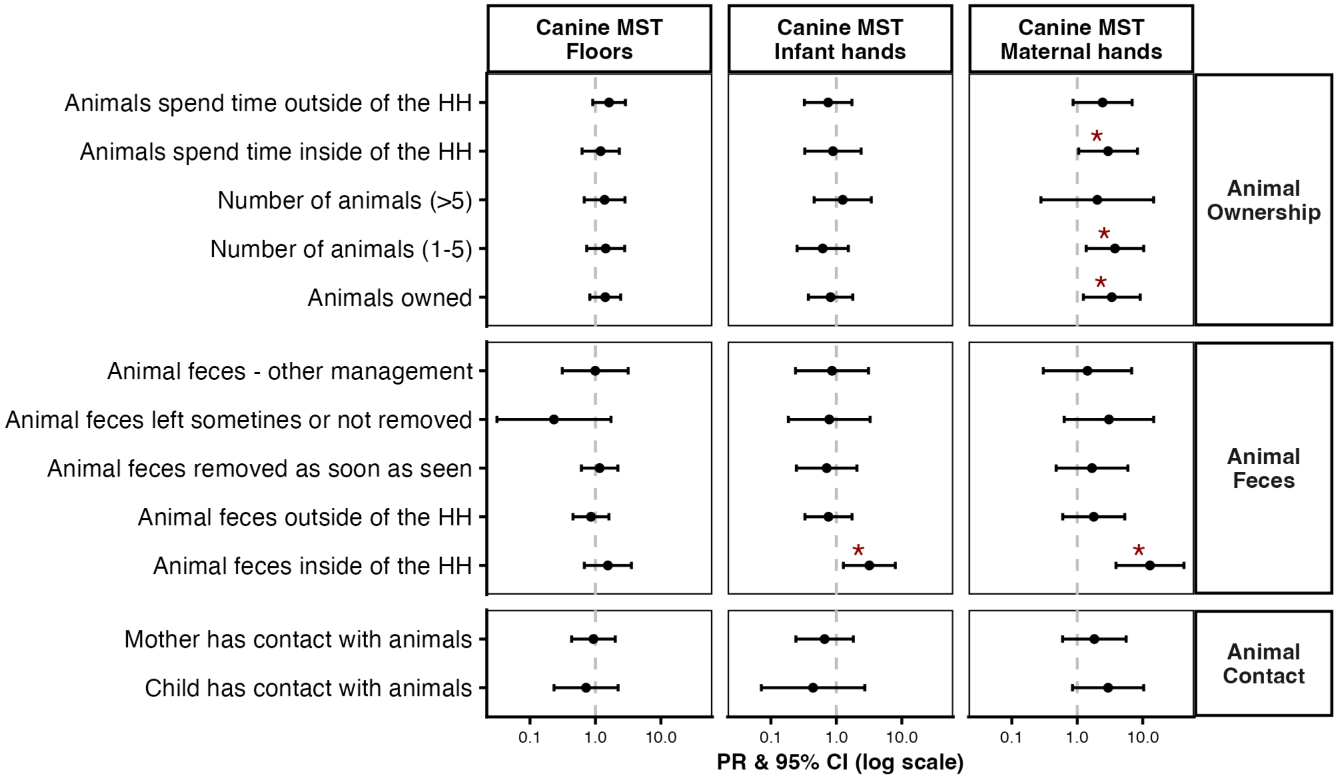


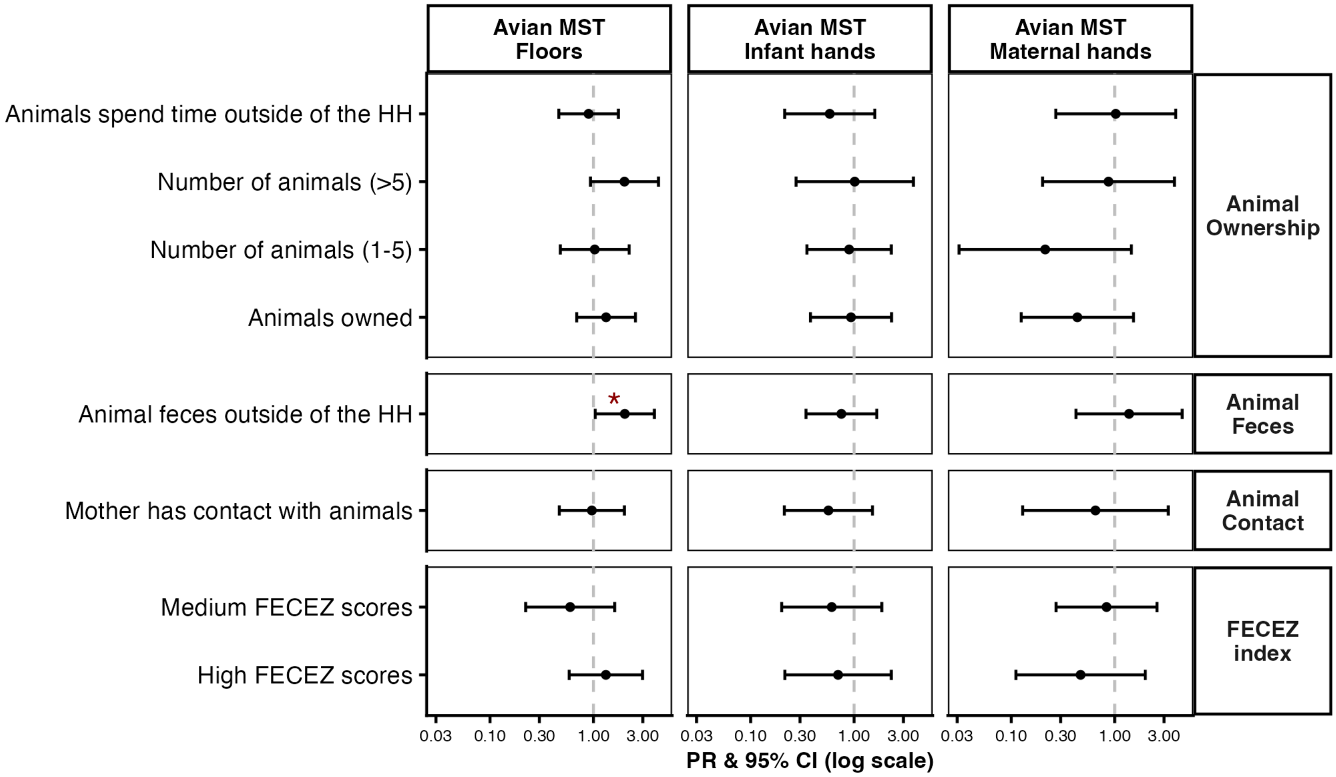

**Figure S4:** Canine and Avian MST markers (abundance) associations

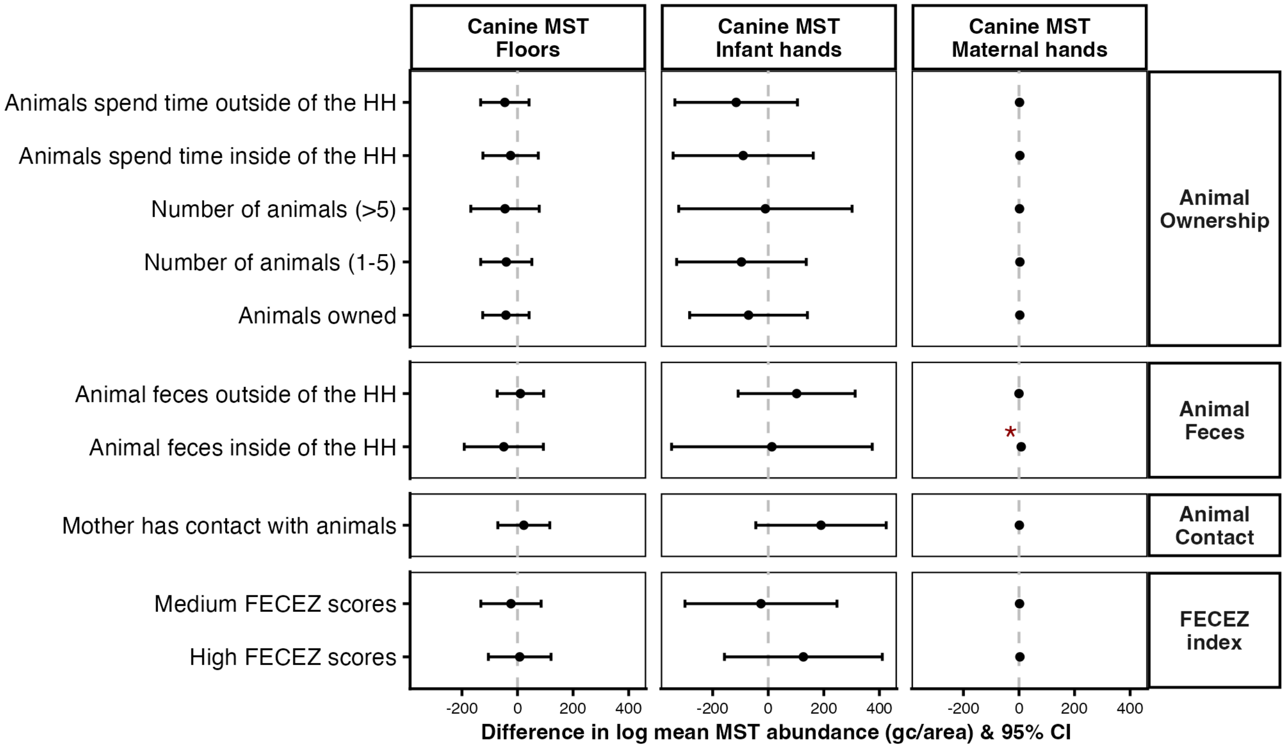


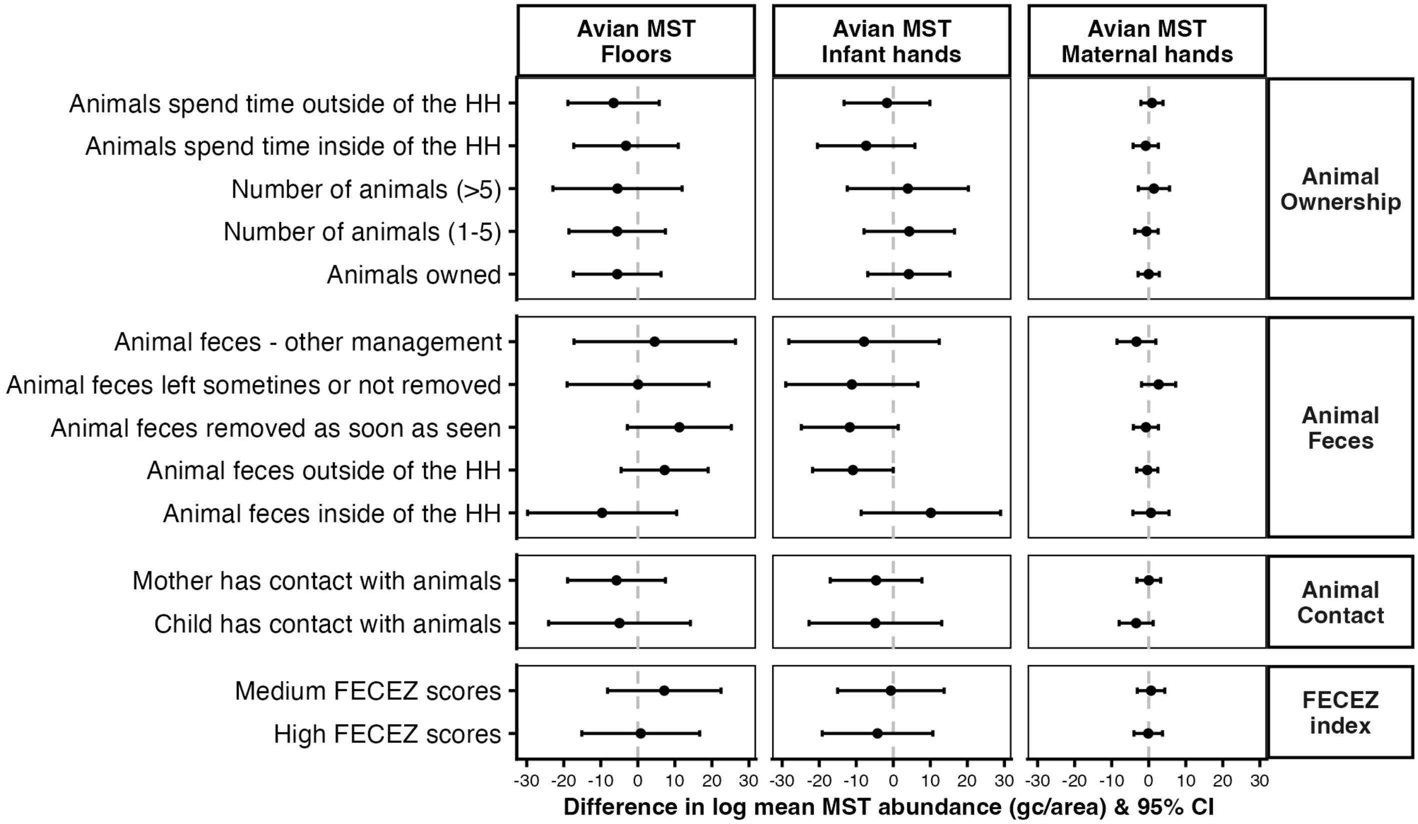
